# Multiplex SARS-CoV-2 Genotyping PCR for Population-Level Variant Screening and Epidemiologic Surveillance

**DOI:** 10.1101/2021.04.20.21255480

**Authors:** Hannah Wang, Jacob A. Miller, Michelle Verghese, Mamdouh Sibai, Daniel Solis, Kenji O. Mfuh, Becky Jiang, Naomi Iwai, Marilyn Mar, ChunHong Huang, Fumiko Yamamoto, Malaya K. Sahoo, James Zehnder, Benjamin A. Pinsky

**Author notes:** **Corresponding Author**: Benjamin A. Pinsky, MD, PhD, 3375 Hillview, Room 2913, Palo Alto, CA 94304. **Alternate Corresponding Author**, Hannah Wang, MD, 3375 Hillview, Room 2913, Palo Alto, CA 94304. Authors HW and JAM contributed equally to this manuscript.

## Abstract

**Background:** Emergence of SARS-CoV-2 variants with concerning phenotypic mutations is of public health interest. Genomic surveillance is an important tool for pandemic response, but many laboratories do not have the resources to support population-level sequencing. We hypothesized that a spike genotyping nucleic acid amplification test (NAAT) could facilitate high-throughput variant surveillance.

**Methods:** We designed and analytically validated a one-step multiplex allele-specific reverse transcriptase polymerase chain reaction (RT-qPCR) to detect three non-synonymous spike protein mutations (L452R, E484K, N501Y). Assay specificity was validated with next-generation whole-genome sequencing. We then screened a large cohort of SARS-CoV-2 positive specimens from our San Francisco Bay Area population.

**Results:** Between December 1, 2020 and March 1, 2021, we screened 4,049 unique infections by genotyping RT-qPCR, with an assay failure rate of 2.8%. We detected 1,567 L452R mutations (38.7%), 34 N501Y mutations (0.84%), 22 E484K mutations (0.54%), and 3 (0.07%) E484K+N501Y mutations. The assay had near-perfect (98-100%) concordance with whole-genome sequencing in a validation subset of 229 specimens, and detected B.1.1.7, B.1.351, B.1.427, B.1.429, B.1.526, and P.2 variants, among others. The assay revealed rapid emergence of L452R in our population, with a prevalence of 24.8% in December 2020 that increased to 62.5% in March 2021.

**Conclusions:** We developed and clinically implemented a genotyping RT-qPCR to conduct high-throughput SARS-CoV-2 variant screening. This approach can be adapted for emerging mutations and immediately implemented in laboratories already performing NAAT worldwide using existing equipment, personnel, and extracted nucleic acid.

**Summary / Key Points:** Emergence of SARS-CoV-2 variants with concerning phenotypes is of public health interest. We developed a multiplex genotyping RT-qPCR to rapidly detect L452R, E484K, and N501Y with high sequencing concordance. This high-throughput alternative to resource-intensive sequencing enabled surveillance of L452R emergence.

## INTRODUCTION

High transmission rates during the coronavirus disease 2019 (COVID-19) pandemic have facilitated the emergence of severe acute respiratory syndrome coronavirus 2 (SARS-CoV-2) variants. The underlying genomic diversity defining these variants is indicative of natural selection of mutations with phenotypic significance.^1–3^ In particular, variants associated with increased transmission, virulence, or resistance to host, vaccine, or antiviral immunity are of significant public health concern.^4–17^ Other mutations could also potentially impact diagnostic sensitivity.^18^ Genomic surveillance that is representative of the newly-infected population is therefore an important tool for pandemic response.

As of March 2021, the United States Centers for Disease Control and Prevention (CDC), European Centre for Disease Prevention and Control (ECDC), and World Health Organization (WHO) identified multiple SARS-CoV-2 lineages as variants of concern or variants of interest (VOC/VOI).^19–21^ Many share the spike protein mutations E484K (B.1.351, B.1.525, B.1.526, P.1, P.2), N501Y (B.1.1.7, B.1.351, P.1), and/or L452R (B.1.427, B.1.429), among others. Such variants have primarily been identified by targeted or whole-genome sequencing. However, due to the required resources, technical expertise, and equipment for sequencing, few public health systems have successfully coordinated timely large-scale sequencing efforts. Among individual institutions that have developed sequencing programs, there is tremendous variability in throughput, selection criteria, and methodology that confound accurate estimates of individual variant prevalence. Because the vast majority of laboratories worldwide do not have the resources required to develop and support large-scale sequencing initiatives, a complementary surveillance strategy is desirable.

Upper respiratory nucleic acid amplification tests (NAATs) are the mainstay of SARS-CoV-2 diagnosis. In contrast to sequencing, nearly all laboratories offering NAATs have equipment and trained personnel for nucleic acid extraction and real-time polymerase chain reaction. We therefore aimed to develop a highly-accessible spike genotyping NAAT for high-throughput and rapid screening of positive SARS-CoV-2 samples. We hypothesized that this assay would facilitate large-scale VOC/VOI screening and track emergence of mutations in our population. This approach enables population-based triaging of samples to centers performing sequencing analysis for lineage determination and increases access to variant surveillance.

## METHODS

### Assay Design

In January 2021, the B.1.1.7, B.1.351, B.1.1.28 descendant P.1, B.1.1.28 descendant P.2, and B.1.427/B.1.429 variant lineages were of primary global and local epidemiologic significance. Informed by recent studies describing mutation phenotypic significance and convergent evolution, we elected to begin screening our population for the spike N501Y (B.1.1.7, B.1.351, P.1), E484K (B.1.351, P.1, P.2), and L452R (B.1.427/B.1.429) mutations.

We therefore designed a one-step multiplex allele-specific reverse transcriptase quantitative polymerase chain reaction (RT-qPCR) to detect these three non-synonymous single nucleotide variants in the spike gene (N501Y: RefSeq NC_045512 23063A>T; E484K: 23012G>A, L452R: 22917 T>G). We selected the wild-type N501 allele (23063A) to serve as a control for those samples that lack any of the targeted mutations.

To permit single reaction multiplexing, we designed primer sets and dual-labeled hydrolysis probes to maximize each probe’s mismatch-ΔTm while maintaining probe specificity and a sufficiently high annealing temperature (Supplementary Table 1). This 2-7°C decrease in each probe’s annealing temperature (ΔTm) when binding SARS-CoV-2 cDNA with a single nucleotide mismatch distinguishes mutant from wild-type nucleic acid templates. Given the proximity between the E484K and N501Y mutations, two primer sets (flanking L452R and E484K/N501Y, respectively) were designed with annealing temperatures similar to or above probe annealing temperatures.

Recognizing the potential for mutations in the targets, on January 26, 2021 we downloaded all available SARS-CoV-2 sequences from GISAID annotated as B.1.1.7 (n=7,864), B.1.351 (n=341), or B.1.427/B.1.429 variants (n=622), as well as all available sequences from NCBI collected prior to December 2020 (n=31,016).^22^ Primer sequences were conserved in >98.8% of all sequences; mutant probe sequences were conserved in >99.7% of variant-specific sequences and were present in only 0.1% of pre-December 2020 NCBI sequences. The wild-type N501 probe sequence was conserved in 99.2% of pre-December 2020 NCBI sequences.

Additional details regarding synthetic controls, oligonucleotides, cycling conditions, fluorescence thresholds, analytical validation, and assay interpretation for reporting are provided in the Supplementary Methods, Supplementary Tables 1-6, and Supplemental Figure 1.

### Clinical Specimens

This study included upper respiratory swab specimens collected between December 1, 2020 and March 3, 2021 from patients as part of routine clinical care. All specimens testing positive for SARS-CoV-2 by NAAT with RT-qPCR C_t_ ≤ 30 or transcription-mediated amplification relative light units (RLU) ≥ 1,100 during this period were subject to multiplex allele-specific genotyping RT-qPCR. All tests were conducted at the Stanford Clinical Virology Laboratory, which serves tertiary-care academic hospitals and affiliated outpatient facilities in the San Francisco Bay Area. After development and analytical validation of our genotyping RT-qPCR, we retrospectively screened all eligible positive specimens dating back to December 1, 2020, and then began prospective genotyping on February 19, 2021. This study was conducted with Stanford institutional review board approval (protocol 48973), and individual consent was waived.

Prior to genotyping RT-qPCR, initial respiratory SARS-CoV-2 NAAT was conducted on a variety of platforms (Table 1, Supplementary Methods).^23–25^ All assays were conducted according to manufacturer and emergency authorization instructions.^26,27^

**Table 1.**
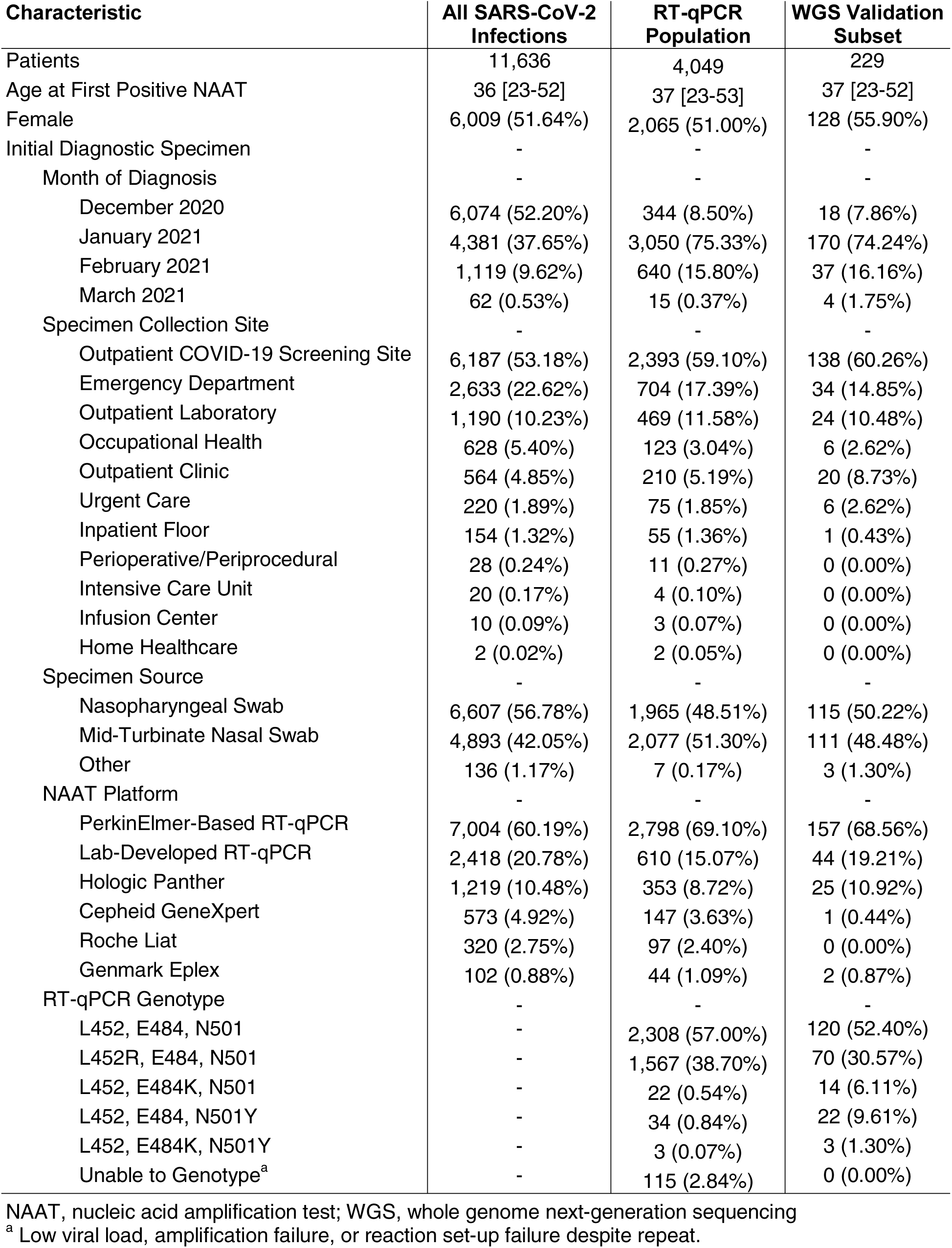
Patient Characteristics and Aggregate Genotyping RT-qPCR Screening Results During Study Period

| Characteristic | All SARS-CoV-2 Infections | RT-qPCR Population | WGS Validation Subset |
| --- | --- | --- | --- |
| Patients | 11,636 | 4,049 | 229 |
| Age at First Positive NAAT | 36 [23-52] | 37 [23-53] | 37 [23-52] |
| Female | 6,009 (51.64%) | 2,065 (51.00%) | 128 (55.90%) |
| Initial Diagnostic Specimen | - | - | - |
| Month of Diagnosis | - | - | - |
| December 2020 | 6,074 (52.20%) | 344 (8.50%) | 18 (7.86%) |
| January 2021 | 4,381 (37.65%) | 3,050 (75.33%) | 170 (74.24%) |
| February 2021 | 1,119 (9.62%) | 640 (15.80%) | 37 (16.16%) |
| March 2021 | 62 (0.53%) | 15 (0.37%) | 4 (1.75%) |
| Specimen Collection Site | - | - | - |
| Outpatient COVID-19 Screening Site | 6,187 (53.18%) | 2,393 (59.10%) | 138 (60.26%) |
| Emergency Department | 2,633 (22.62%) | 704 (17.39%) | 34 (14.85%) |
| Outpatient Laboratory | 1,190 (10.23%) | 469 (11.58%) | 24 (10.48%) |
| Occupational Health | 628 (5.40%) | 123 (3.04%) | 6 (2.62%) |
| Outpatient Clinic | 564 (4.85%) | 210 (5.19%) | 20 (8.73%) |
| Urgent Care | 220 (1.89%) | 75 (1.85%) | 6 (2.62%) |
| Inpatient Floor | 154 (1.32%) | 55 (1.36%) | 1 (0.43%) |
| Perioperative/Periprocedural | 28 (0.24%) | 11 (0.27%) | 0 (0.00%) |
| Intensive Care Unit | 20 (0.17%) | 4 (0.10%) | 0 (0.00%) |
| Infusion Center | 10 (0.09%) | 3 (0.07%) | 0 (0.00%) |
| Home Healthcare | 2 (0.02%) | 2 (0.05%) | 0 (0.00%) |
| Specimen Source | - | - | - |
| Nasopharyngeal Swab | 6,607 (56.78%) | 1,965 (48.51%) | 115 (50.22%) |
| Mid-Turbinate Nasal Swab | 4,893 (42.05%) | 2,077 (51.30%) | 111 (48.48%) |
| Other | 136 (1.17%) | 7 (0.17%) | 3 (1.30%) |
| NAAT Platform | - | - | - |
| PerkinElmer-Based RT-qPCR | 7,004 (60.19%) | 2,798 (69.10%) | 157 (68.56%) |
| Lab-Developed RT-qPCR | 2,418 (20.78%) | 610 (15.07%) | 44 (19.21%) |
| Hologic Panther | 1,219 (10.48%) | 353 (8.72%) | 25 (10.92%) |
| Cepheid GeneXpert | 573 (4.92%) | 147 (3.63%) | 1 (0.44%) |
| Roche Liat | 320 (2.75%) | 97 (2.40%) | 0 (0.00%) |
| Genmark Eplex | 102 (0.88%) | 44 (1.09%) | 2 (0.87%) |
| RT-qPCR Genotype | - | - | - |
| L452, E484, N501 | - | 2,308 (57.00%) | 120 (52.40%) |
| L452R, E484, N501 | - | 1,567 (38.70%) | 70 (30.57%) |
| L452, E484K, N501 | - | 22 (0.54%) | 14 (6.11%) |
| L452, E484, N501Y | - | 34 (0.84%) | 22 (9.61%) |
| L452, E484K, N501Y | - | 3 (0.07%) | 3 (1.30%) |
| Unable to Genotype<sup>a</sup> | - | 115 (2.84%) | 0 (0.00%) |

NAAT, nucleic acid amplification test; WGS, whole genome next-generation sequencing
<sup>a</sup> Low viral load, amplification failure, or reaction set-up failure despite repeat.

### Assay Validation with Next-Generation Sequencing

A subset of wild-type and mutant specimens genotyped by RT-qPCR were confirmed by WGS in a validation dataset. We adapted an existing WGS pipeline for poliovirus genotyping to conduct SARS-CoV-2 whole-genome amplicon-based sequencing (Supplementary Methods).^28–30^ Pangolin was used for PANGO lineage assignment, while Nextclade v0.14.2 and auspice.us 0.5.0. were used to assign Nextstrain clades and perform phylogenetic placement.^2,30,31^ PANGO lineage names and Nextstrain clade names were assigned as of April 5, 2021.

### Statistical Analysis

Baseline continuous data were compared with t-tests or Wilcoxon rank-sum tests, whereas categorical data were compared with Fisher exact tests. Positive percent agreement (PPA) and negative percent agreement (NPA) were reported with Clopper-Pearson score 95% binomial confidence intervals using next-generation sequencing as the reference method. Analyses were conducted using the R statistical software package. This study was reported in accordance with STARD guidelines.^32^

## RESULTS

### Population-Level Screening Results

During the period between December 1, 2020 and March 3, 2021, our clinical virology laboratory reported 165,501 negative and 13,083 positive SARS-CoV-2 NAATs among 119,154 unique patients. These 13,083 positive NAATs corresponded to 11,635 patients (Table 1). We assumed this reflected approximately 11,635 unique infections, presuming no re-infection during the study period.

After triaging specimens with higher viral load and sufficient remaining material, we screened 4,049 specimens from unique patients by genotyping RT-qPCR (Table 1, Figure 1). Amplification curves from RT-qPCR were interpreted according to Supplementary Table 4, with example cases presented in Supplementary Figure 2. A total of 174 specimens had amplification failure on initial RT-qPCR, either due to low viral load, amplification failure, or reaction setup error. Among these specimens, 59 were successfully genotyped on repeat RT-qPCR and are included in the analysis dataset, while the remaining 115 were unable to genotype, corresponding to an assay failure rate of 2.8%. Among initial diagnostic specimens tested by RT-qPCR with available C_t_ values (3,142), median viral load was significantly lower among samples unable to genotype (C_t_ 28.8 vs 18.8, *p<*0.001).

**Figure 1.** Flow chart of specimens sent for nucleic acid amplification tests (NAAT) during the study period and number of specimens ultimately genotyped by multiplex RT-qPCR. *Predominantly specimens with insufficient material, low viral load (C_t_>30, RLU<1,100) or positive specimens in December prior to universal institution-wide genotyping.

Among the 3,934 genotyped specimens, we identified 1,567 (38.7%) with isolated L452R mutations, 34 (0.84%) with isolated N501Y mutations, 22 (0.54%) with isolated E484K mutations, and 3 (0.07%) with E484K+N501Y mutations.

### Assay Validation with Next-Generation Sequencing

In addition to validating the assay’s analytical performance (Supplementary Tables 5-6), we assessed clinical positive (PPA) and negative percent agreement (NPA) of genotyping RT-qPCR using WGS as a reference method. Among 229 specimens from unique patients assessed by both RT-qPCR and WGS (Table 1), PPA of RT-qPCR for L452R, E484K, and N501Y were: 100% (95% CI 95-100%), 100% (95% CI 80-100%), and 100% (95% CI 86-100%), respectively (Table 2). There were no false negatives by RT-qPCR for L452R, E484K, or N501Y. Furthermore, there were no false positives for E484K (NPA: 100%, 95% CI 98-100%) or N501Y (NPA: 100%, 95% CI 98-100%).

**Table 2.** Genotyping RT-qPCR Validation Against Whole-Genome Next-Generation Sequencing

| RT-qPCR | Whole-Genome Next-Generation Sequencing |  |  |  |  |  |
| --- | --- | --- | --- | --- | --- | --- |
|  | L452R |  | E484K |  | N501Y |  |
|  | Detected | Not Detected | Detected | Not Detected | Detected | Not Detected |
| Detected | 67 | 3<sup>a</sup> | 17 | 0 | 25 | 0 |
| Not Detected | 0 | 159 | 0 | 212 | 0 | 204 |
| PPA (95% CI)<sup>b</sup> | 100% (95-100%) | - | 100% (80-100%) | - | 100% (86-100%) | - |
| NPA (95% CI)<sup>b</sup> | - | 98% (95-99%) | - | 100% (98-100%) | - | 100% (98-100%) |

CI, confidence interval; PPA, positive percent agreement; NPA, negative percent agreement. <sup>a</sup> Three specimens positive for L452R by RT-qPCR but negative for L452R by sequencing. Further characterization of these samples provided in results. <sup>b</sup> RT-qPCR relative to next-generation sequencing.

There were three specimens positive for L452R by RT-qPCR but negative for by WGS (NPA: 98%, 95% CI 95-99%). These had L452R C_t_ values of 20.0, 22.8, and 31.2 with appropriate amplification curves and appropriate N501 wild-type (Cy3.5) internal control amplification (C_t_ 21.0, 18.4, 30.0). Sequencing coverage and codon calls at the L452 loci were 1,890/1,907 (99%), 2,407/2,424 (99%), and 2,817/2,841 (99%) for these three specimens, which were assigned lineages of B.1.1.222, B.1.2, and B.1.243, respectively. All three specimens were re-extracted and repeated by RT-qPCR with the same result.

Among the 229 specimens tested by both RT-qPCR and WGS, the distribution of PANGO lineages is presented in Table 3, and a phylogenetic tree of Nextstrain clades is presented in Figure 2. The 120 specimens which were wild-type by RT-qPCR were most commonly assigned lineages of B.1.2 (34%) and B.1 (12%). Among the 70 specimens with isolated L452 mutations by RT-qPCR, 43 (60%) and 23 (33%) were B.1.427 and B.1.429, respectively. Among the 22 specimens with isolated N501Y mutations by RT-qPCR, 17 (77%) were B.1.1.7. Among the 14 specimens with isolated E484K mutations by RT-qPCR, 11 (79%) were P.2, 2 (14%) were B.1, and 1 (7%) was B.1.526 (Figure 3). The three specimens with E484K+N501Y mutations were all of B.1.351 lineage. Sequencing quality measures and GISAID accessions are provided in Supplementary Tables 7-8.

**Figure 2.** Nextclade phylogenetic tree of 2,130 SARS-CoV-2 genomes, including 225 of the 229 sequenced genomes from this study, and 1,905 genomes from the Nextstrain global reference tree (left panel). There were 3 sequenced genomes excluded due to discordance between PCR and sequencing, and 1 genome excluded due to insufficient quality to assign a clade. The 225 included genomes are colored by RT-qPCR genotyping result, with each circle representing a sequenced genome. Branch length corresponds to nucleotide divergence. Arrowheads point to mutations identified either sporadically, or in rare lineages. Isolates with N501Y mutations predominantly belong to clade 20I/501Y.V1 and lineage B.1.1.7, though sporadic mutations are noted in clade 20G lineage B.1.2 (top three green arrowheads) and clade 20A lineage B.1.480 (bottom green arrowhead). In addition to the clade 20B lineage P.2 variant of concern, E484K RT-qPCR also identified a clade 20C lineage B.1.526 variant of interest (top two yellow arrowheads), as well as a clade 20A proposed lineage B.1 (bottom yellow arrowhead). Isolates with L452R mutation (turquoise) belong predominantly to clade 20C, and lineages B.1.429 and B.1.427 (right panel). Sporadic L452R mutation was noted within one sample belonging to clade 20A lineage B.1 (turquoise arrowhead). PANGO lineage names and Nextstrain clade names were assigned as of April 5, 2021.

**Figure 3.** Nextclade phylogenetic tree from Figure 2 magnified to visualize isolates with E484K mutations in clade 20B lineage P.2 (left panel), and isolates with combined N501Y+E484K mutations, all of which belong to clade 20H/501Y.V2 lineage B.1.351 (right panel). They are shown next to their nearest neighbors from the Nextstrain global reference tree, with branch length corresponding to nucleotide divergence. PANGO lineage names and Nextstrain clade names were assigned as of April 5, 2021.

**Table 3.** PANGO Lineage Distribution of Screen-Detected Variants by RT-qPCR

| PANGO Lineage | RT-qPCR Genotype |  |  |  |  |
| --- | --- | --- | --- | --- | --- |
|  | L452R, E484, N501 | L452, E484K, N501 | L452, E484, N501Y | L452, E484K, N501Y | L452, E484, N501 |
| B.1 | 2 (3) | 2 (14) | 1 (5) | 0 (0) | 14 (12) |
| B.1.1 | 0 (0) | 0 (0) | 0 (0) | 0 (0) | 2 (2) |
| B.1.1.174 | 0 (0) | 0 (0) | 0 (0) | 0 (0) | 1 (1) |
| B.1.1.222 | 1 (1)<sup>a</sup> | 0 (0) | 0 (0) | 0 (0) | 1 (1) |
| B.1.1.228 | 0 (0) | 0 (0) | 0 (0) | 0 (0) | 1 (1) |
| P.2<sup>b</sup> | 0 (0) | 11 (79) | 0 (0) | 0 (0) | 0 (0) |
| B.1.1.291 | 0 (0) | 0 (0) | 0 (0) | 0 (0) | 1 (1) |
| B.1.1.368 | 0 (0) | 0 (0) | 0 (0) | 0 (0) | 2 (2) |
| B.1.1.416 | 0 (0) | 0 (0) | 0 (0) | 0 (0) | 3 (3) |
| B.1.1.432 | 0 (0) | 0 (0) | 0 (0) | 0 (0) | 1 (1) |
| B.1.1.517 | 0 (0) | 0 (0) | 0 (0) | 0 (0) | 1 (1) |
| B.1.1.519 | 0 (0) | 0 (0) | 0 (0) | 0 (0) | 8 (7) |
| B.1.1.7<sup>c</sup> | 0 (0) | 0 (0) | 17 (77) | 0 (0) | 0 (0) |
| B.1.139 | 0 (0) | 0 (0) | 0 (0) | 0 (0) | 2 (2) |
| B.1.2 | 1 (1)<sup>a</sup> | 0 (0) | 3 (14) | 0 (0) | 41 (34) |
| B.1.232 | 0 (0) | 0 (0) | 0 (0) | 0 (0) | 5 (4) |
| B.1.234 | 0 (0) | 0 (0) | 0 (0) | 0 (0) | 3 (3) |
| B.1.239 | 0 (0) | 0 (0) | 0 (0) | 0 (0) | 1 (1) |
| B.1.240 | 0 (0) | 0 (0) | 0 (0) | 0 (0) | 3 (3) |
| B.1.243 | 1 (1)<sup>a</sup> | 0 (0) | 0 (0) | 0 (0) | 6 (5) |
| B.1.265 | 0 (0) | 0 (0) | 0 (0) | 0 (0) | 1 (1) |
| B.1.311 | 0 (0) | 0 (0) | 0 (0) | 0 (0) | 6 (5) |
| B.1.323 | 0 (0) | 0 (0) | 0 (0) | 0 (0) | 1 (1) |
| B.1.351<sup>c</sup> | 0 (0) | 0 (0) | 0 (0) | 3 (100) | 0 (0) |
| B.1.361 | 0 (0) | 0 (0) | 0 (0) | 0 (0) | 1 (1) |
| B.1.369 | 0 (0) | 0 (0) | 0 (0) | 0 (0) | 2 (2) |
| B.1.375 | 0 (0) | 0 (0) | 0 (0) | 0 (0) | 2 (2) |
| B.1.427<sup>c</sup> | 42 (60) | 0 (0) | 0 (0) | 0 (0) | 0 (0) |
| B.1.429<sup>c</sup> | 23 (33) | 0 (0) | 0 (0) | 0 (0) | 0 (0) |
| B.1.453 | 0 (0) | 0 (0) | 0 (0) | 0 (0) | 1 (1) |
| B.1.480 | 0 (0) | 0 (0) | 1 (5) | 0 (0) | 0 (0) |
| B.1.526<sup>b</sup> | 0 (0) | 1 (7) | 0 (0) | 0 (0) | 0 (0) |
| B.1.561 | 0 (0) | 0 (0) | 0 (0) | 0 (0) | 4 (3) |
| B.1.565 | 0 (0) | 0 (0) | 0 (0) | 0 (0) | 1 (1) |
| B.1.568 | 0 (0) | 0 (0) | 0 (0) | 0 (0) | 1 (1) |
| B.1.577 | 0 (0) | 0 (0) | 0 (0) | 0 (0) | 1 (1) |
| B.1.596 | 0 (0) | 0 (0) | 0 (0) | 0 (0) | 2 (2) |
| B.1.599 | 0 (0) | 0 (0) | 0 (0) | 0 (0) | 1 (1) |
| **Total** | **70** | **14** | **22** | **3** | **120** |

Data presented as n (column %) <sup>a</sup> Three specimens positive for L452R by RT-qPCR but negative for L452R by next-generation sequencing. <sup>b</sup> CDC-classified variants of interest <sup>c</sup> CDC-classified variants of concern

### Epidemiologic Emergence of L452R

During the study period, we observed rapid emergence of the L452R mutation in our population (Figure 4, Table 4). The majority of such specimens assessed by WGS were B.1.427/B.1.429 variants (93%). Although the number of new infections in our population decreased between January and March 2021, the proportion of new infections harboring the L452R mutation increased from 24.8% in December 2020 to 62.5% in March 2021.

**Figure 4.** Local epidemiologic emergence of L452R mutations in our genotyped population of 3,934 from December 2020 to March 2021. Bar plot and left y-axis represent number of new unique genotyped infections by week grouped by RT-qPCR-defined genotype. Scatter plot and right y-axis represents proportion of new unique genotyped infections with L452R genotype, with 95% Clopper-Pearson confidence interval.

**Table 4.** Local Epidemiologic Emergence of L452R

| Week of Year, 2020-2021 | Total New Infections Genotyped<sup>a</sup> | L452R Infections |
| --- | --- | --- |
| Weeks 49-52: 2020-12-01 to 2020-12-31<sup>b</sup> | 476 | 118 (24.8%) |
| Week 1: 2021-01-01 | 905 | 300 (33.1%) |
| Week 2: 2021-01-08 | 888 | 330 (37.2%) |
| Week 3: 2021-01-15 | 598 | 267 (44.6%) |
| Week 4: 2021-01-22 | 329 | 141 (42.9%) |
| Week 5: 2021-01-29 | 265 | 145 (54.7%) |
| Week 6: 2021-02-05 | 199 | 102 (51.3%) |
| Week 7: 2021-02-12 | 120 | 70 (58.3%) |
| Week 8: 2021-02-19 | 106 | 64 (60.4%) |
| Week 9: 2021-02-26 | 48 | 30 (62.5%) |
| **Total** | 3,934 | 1,567 (39.8%) |

<sup>a</sup> Confirmed by nucleic acid amplification testing, with diagnosis date assigned by first positive test date.
<sup>b</sup> Grouped due to fewer tested specimens in early December 2020.
<sup>c</sup> Proportion of cases in each week positive for L452R.

## DISCUSSION

Rapid, accessible, and high-throughput epidemiologic surveillance for SARS-CoV-2 variants is of public health interest. We describe a one-step multiplex RT-qPCR assay that facilitated screening of approximately 4,000 unique infections during a period of high transmission in the San Francisco Bay Area. This assay had excellent concordance with WGS and revealed rapid emergence of variants harboring L452R. In contrast to WGS, the widely-available equipment, resources, and trained personnel make this RT-qPCR assay accessible to nearly all laboratories already offering SARS-CoV-2 NAATs. All current CDC-classified variants of concern (B.1.1.7, P.1, B.1.351, B.1.427, B.1.429) and most variants of interest (P.2, B.1.525, some but not all B.1.526) harbor at least one of the L452R, E484K, or N501Y spike protein mutations, enabling population-level triaging by RT-qPCR to centers performing WGS for lineage determination. This approach has implications for outbreak surveillance, diagnostic testing, transmissibility, virulence, and vaccine and/or antiviral efficacy.

The CDC, ECDC, and WHO have identified multiple SARS-CoV-2 lineages as variants of concern (VOC) or variants of interest (VOI).^19–21^ Many of these variants share the E484K mutation (B.1.351, B.1.525, some B.1.526, P.1, P.2), N501Y mutation (B.1.1.7, B.1.351, P.1), and/or the L452R mutation (B.1.427, B.1.429). Previously-reported studies guided by large-scale genomic surveillance have suggested increased transmissibility, virulence, or reduced vaccine/antiviral efficacy/neutralization corresponding to the phenotypic significance of these spike protein mutations.^4–17^ Moreover, mutations in the spike gene (*S* gene) may lead to false negative NAAT results.^18^

While whole-genome sequencing is required to confirm infection with a specific variant, not all centers have the required resources or personnel to conduct large-scale variant genotyping on a high proportion of incident cases. Allele-specific NAAT-based methods are an alternative to large-scale sequencing programs, but few methods have been described. Korukluoglu et al. recently described a one-step RT-qPCR assay to detect the N501Y and HVdel69-70 using allele-specific forward primers, reserving ORF1ab as an internal control.^33^ This assay is conducted in two separate reactions, with the first targeting N501, ORF1ab, and HV69-70del, while the second amplifies N501Y, ORF1ab, and HV69-70del. The authors described high concordance (100%) with Sanger and next-generation sequencing. A disadvantage of this method is the absence of screening for E484K and the requirement for two separate reactions. More recently, Banada et al. reported an RT-qPCR method for detection of N501Y using melt curve analysis.^34^ This assay includes allele-specific molecular beacon probes for N501 and N501Y conjugated to different fluorophores, similar to our assay. Twenty patient specimens were tested and a subset were confirmed by Sanger sequencing.

Vogels et al. have also described an RT-qPCR assay to detect the ORF1ab 3675-3677 and spike HV69-70 deletions.^35^ The deletions are detected via ORT1ab-Cy5 and spike-HEX probe dropout, reserving the CDC N1 primer/probe set as an internal control. This assay was concordant with 76 sequenced specimens. Limitations of this assay, as described by the authors, are N1-FAM autofluorescence leading to false B.1.1.7 drop-out and absence of the ORF1ab 3675-3677 deletion in select B.1.351 isolates. While these deletions are surrogates for B.1.1.7, B.1.351, and P.1, screening for N501Y and E484K may be more specific to variants of concern/interest and appear to be causally-associated with concerning phenotypes. Another genotyping NAAT utilized a two-step approach combining the CDC-based laboratory-developed RT-qPCR and the ThermoFisher TaqPath COVID-19 RT-PCR.^36^ With the intent to identify B.1.1.7 variants, the authors screened 1,035 samples for TaqPath *S* gene dropout and genotyped two specimens by multiplex droplet digital PCR amplifying HV69-70del, N501Y, del145, and S982A in two reactions, which were positive for these four mutations. Next-generation sequencing confirmed B.1.1.7 lineage. Limitations of this genotyping approach include dependence upon *S* gene dropout followed by more labor-intensive ddPCR.

The largest NAAT-based genotyping series thus far was a national effort in France recently reported by Haim-Boukobza et al.^37^ The authors used two separate assays to screen for the HVdel69-70 and N501Y mutations in 35,208 samples, with an uninterpretable assay rate of 19.0% and sequencing concordance Kappa coefficient of 0.87-0.88.

To our knowledge, the present study is the largest NAAT-based SARS-CoV-2 genotyping initiative in the United States. While large multi-institutional programs such as the United Kingdom COVID-19 genomics consortium have sequenced more than 250,000 specimens, many public health systems or private institutions are unable to conduct high-throughput genotyping due to the required cost, equipment, and expertise. The RT-qPCR assay we describe is accessible to any laboratory conducting RT-qPCR SARS-CoV-2 testing, and shares much of the same equipment and reagents. Specimens positive for SARS-CoV-2 by initial NAAT can be rapidly genotyped within hours using the same extracted nucleic acid, and the cost of this assay’s custom oligonucleotides are less than one U.S. Dollar per reaction. As proof of concept, we have validated this assay on various qPCR instruments and master mixes, demonstrating that individual laboratories may adapt this assay to existing NAAT workflows.

As evidence of the feasibility of high-throughput testing, we rapidly implemented reflex genotyping RT-qPCR for positive specimens and screened more than 4,000 infections, thereby facilitating local surveillance of L452R mutation emergence. Our results are consistent with a large-scale whole-genome sequencing initiative of 2,172 specimens from 44 counties in California.^12^ This series suggested a 19-24% increase in transmissibility relative to wild-type circulating strains, and was the basis for L452R surveillance in our study. Due to the local prevalence of circulating L452R mutations, our screening initiative identified approximately 1,500 unique infections harboring L452R mutations.

This assay has several limitations. Sporadic isolated L452R, E484K, and N501Y mutations are well-described and are not necessarily indicative of VOC/VOI. For example, in our series of 229 sequenced specimens genotyped by RT-qPCR, isolated E484K or N501Y mutations by PCR were VOC/VOI in 86% and 77% of cases, respectively, while the remaining were uncommon variants. Although our assay does not screen for several other mutations of interest (HVdel69-70, K417N, P681H, Y453F), it is advantageous that such variants are screen-detected via N501Y (B.1.1.7: del69-70, P681H), E484K (P.1 and B.1.351: K417N), and/or L452R. An additional limitation is the lack of wild-type probes for L452 and E484. Although this choice was intentional to maximize efficiency, it introduces the possibility that synonymous or alternate non-synonymous mutations (e.g. E484Q) within the probe binding site could lead to an erroneous result. Lastly, variants may develop new mutations in primer and/or probe sites, potentially leading to false negatives. As such, these primer/probe sequences should be regularly checked against all available GISAID sequences, and new VOC/VOI should be monitored for potential assay interference. An additional limitation is exclusion of samples with lower viral load in an effort to successfully genotype. Although our assay failure rate was low (2.8%), analytical sensitivity could be improved by increasing template volume. Although the present series of 4,049 infections is a subset of the 11,635 unique consecutive infections between December 2020 and March 2021, the genotyped population was similar to the overall population (Table 1). Nevertheless, our results may not be necessarily generalizable to all new infections.

We developed and rapidly implemented a one-step multiplex allele-specific RT-qPCR assay to conduct rapid and high-throughput SARS-CoV-2 variant screening in the San Francisco Bay Area. The assay had near-perfect concordance with whole-genome next-generation sequencing and facilitates detection of all common and uncommon variants of concern/interest harboring the L452R, E484K, or N501Y mutations. This approach can be adapted for emerging mutations and implemented in laboratories already conducting SARS-CoV-2 NAAT using existing resources and extracted nucleic acid.

## Data Availability

Data referred to in the manuscript is available upon request.

## ACKNOWLEDGEMENTS

We sincerely thank the Stanford Clinical Virology Laboratory staff for their dedication and commitment to patient care in the face of unprecedented challenges presented by the COVID-19 pandemic. We also thank the Stanford Protein and Nucleic Acid Facility for oligonucleotide synthesis.

