## Supplementary Methods for "Multiplex SARS-CoV-2 Genotyping PCR for Population-Level Variant Screening and Epidemiologic Surveillance"

**Running Title:** SARS-CoV-2 Variant Genotyping PCR

**Corresponding Author:	Alternate Corresponding Author:**

Benjamin A. Pinsky, MD, PhD	Hannah Wang, MD

3375 Hillview, Room 2913	3375 Hillview, Room 2913

Palo Alto, CA 94304	Palo Alto, CA 94304

*Assay Design*

We used synthetic whole-genome RNA fragments as a wild-type control (Twist Bioscience, San Francisco, CA, USA) diluted to 10^4^ copies/µL in Tris-EDTA buffer (10 mM Tris, 1 mM EDTA). Due to manufacturing restrictions for SARS-CoV-2-specific dsDNA gene fragments, we pooled six individual ssDNA mutant amplicons (Elim Biopharmaceuticals, Hayward, CA, USA) in equimolar ratios diluted to 10^4^ copies/µL each in Tris-EDTA buffer (10 mM Tris, 1 mM EDTA) (Supplementary Table 2). The two primer sets and three mutant probes used in this assay amplify two of these six amplicons, while the other four were added for other candidate assays. Primers and dual-labeled BHQ-quenched hydrolysis probes were ordered from the Stanford Protein and Nucleic Acid Facility (Supplementary Table 1), rehydrated to 100µM in Tris-EDTA buffer, and mixed to create bulk primer/probe mix.

Primer/probe mix (1 µL) was combined with a one-step RT-qPCR system (12.5 µL master mix + 0.5 µL *Taq* polymerase, SuperScript™ III Platinum™ One-Step qRT-PCR Kit, Invitrogen, Carlsbad), nuclease-free water (6.0 µL), and template (5.0 µL) in a 25µL reaction (Supplementary Table 3). All experiments were conducted on a BioRad CFX96 real-time PCR instrument in 96-well plates (BioRad, Hercules, CA, USA). One mutant control (pooled ssDNA mutant amplicons) and one wild-type control (Twist whole-genome synthetic RNA) were included in each RT-qPCR experiment. Cycling conditions were: 52°C for 15:00, 94°C for 2:00, and then 45 cycles of 94°C for 00:15, 57.0°C for 00:40, and 68°C for 00:20. Annealing temperature was optimized with a temperature gradient.

Fluorescence was collected in all channels (1, N501Y-FAM; 2, L452R-HEX; 3, N501-Cy3.5 (ROX); 4, E484K-Cy5; 5, no probe). Fixed fluorescence thresholds of 500 relative fluorescence units ([RFU], N501Y-FAM), 1000 RFU (L452R-HEX), 50 RFU (N501-Cy3.5), and 200 RFU (E484K-Cy5) were used to determine the threshold cycle (C_t_). Assay interpretation is described in detail in Supplementary Table 4.

*Analytical Performance*

To determine the lower limit of detection (LLOD), the pool of six mutant ssDNA oligonucleotides described above was diluted to 100 copies/µL template, 10 copies/µL, 5 copies/µL, and 1 copies/µL in Tris-EDTA buffer in replicates of 20. Any amplification crossing the fluorescence threshold (Supplementary Table 4) was regarded as detection. The 95% LLOD was determined by fitting these data to a probit regression curve. The respective 95% LLODs for the L452R, E484K, and N501Y targets were 1.5 (95% CI 1.0-3.1), 16.0 (11.1-40.3), and 23.6 (14.0-29.4) copies/µL template (Supplementary Table 5). Assay linearity was assessed from 0.0 to 6.0 log10 copies/µL template (Supplementary Table 6, Supplementary Figure 1). We observed no non-specific N501 wild-type (Cy3.5) amplification even at high (10^6^ copies/µL) mutant ssDNA copy number; similarly, we observed no non-specific L452R, E484K, or N501Y non-specific amplification at high (10^6^ copies/µL) wild-type TWIST synthetic RNA copy number (Supplementary Table 6). Precision was not assessed for this qualitative assay.

*Next-Generation Sequencing Validation*

A subset of wild-type and mutant specimens genotyped by RT-qPCR were confirmed by WGS in a validation dataset. We adapted an existing WGS pipeline for poliovirus genotyping to conduct SARS-CoV-2 whole-genome amplicon-based sequencing.^1–3^

For target enrichment, we designed 28 primersets to generate 28 overlapping amplicons of approximately 1,200 nucleotides each using PrimalSeq.^2^ Even- and odd-numbered primersets were pooled separately, and then the whole genome was amplified in two reactions via long-range PCR using 10 μL NEB Luna 2X buffer, 1 μL NEB enzyme mix, 2 μL nuclease-free water, 2 μL primer pool (50nM each primer), and 5 μL extracted nucleic acid. Long-range PCR conditions were: 52°C for 30:00, 94°C for 2:00, and then 40 cycles of 94°C for 00:15, 55.0°C for 00:30, and 68°C for 02:00 prior to final 68°C extension for 10:00.

Libraries were prepared using NEBNext library preparation reagents for Illumina (New England BioLabs, Ipswich, MA). Products from the two long-range PCR reactions (40 μL) were pooled and purified with 36 μL (0.9X ratio) AMPure XP beads (Beckman Coulter, Brea, CA) and 80% ethanol, then resuspended into 32 μL AVE elution buffer. Purified cDNA was quantified using the Qubit dsDNA broad range assay. Libraries were then fragmented at 37°C for 30:00 using 2 μL 10X fragmentation buffer, 1 μL 200mM MgCl_2_, 2 μL fragmentase, and 15 μL purified cDNA from long-range PCR.

Fragmented libraries (20 μL) were then purified with 36 μL (1.8X ratio) AMPure XP beads and 80% ethanol, then resuspended into 32 μL AVE elution buffer. Purified fragmented cDNA (30 μL) was subject to end-repair/dA tailing using 1.5 μL NEB end-repair enzyme mix and 3.5 μL end-repair reaction buffer (20°C for 30:00, 65°C for 30:00). We ligated single-index NEBNext adapters according to manufacturer recommendations in a 45 μL reaction containing 35 μL end-repaired product. This product was again purified with 0.9X AMPure XP beads and 80% ethanol, then resuspended in 28 μL buffer.

We indexed libraries using NEB single index primers in a 50 μL reaction containing 25 μL NEBNext high-fidelity 2X master mix, 1 μL universal primer, 1 μL index primer, and 23 μL adapter-ligated cDNA. Thermocycler settings for indexing were: 94°C for 00:30, then 12 cycles of 94°C for 00:10, 65.0°C for 00:30, and 72°C for 00:30. Indexed libraries were then purified with 18 μL (0.9X ratio) AMPure XP beads and 80% ethanol, then resuspended into 32 μL AVE elution buffer.

Indexed library fragment size and concentration were measured with a BioAnalyzer 2100, then diluted to 15pM. Each sequencing run contained one no template control, one wild-type TWIST synthetic whole-genome RNA control, and up to 46 clinical specimens. Libraries were sequenced on an Illumina MiSeq using single-end 150-cycle sequencing using the MiSeq reagent kit V3.

Genomes were assembled via a custom assembly and bioinformatics pipeline using NCBI NC_045512.2 as reference. Whole-genome sequences with at least 90% genome coverage to a depth of at least 10 reads were accepted for interpretation. Mutation calling required a depth of at least 12 reads with a minimum variant frequency of 20%. Lineage name was assigned using PANGOLIN.^3^

*Clinical Specimen NAAT Platforms*

Prior to genotyping RT-qPCR, initial respiratory SARS-CoV-2 NAAT was conducted on a variety of platforms (Table 1).^4–6^ These included: 1) a previously-described laboratory-developed reverse transcription quantitative polymerase chain reaction (RT-qPCR) targeting the envelope gene (*E* gene) on the Rotor-Gene Q (Qiagen, Germantown, MD)^4–6^; 2) a laboratory-developed RT-qPCR assay utilizing a PerkinElmer kit targeting the ORF1ab and nucleocapsid gene ([*N* gene] PerkinElmer, San Jose, CA); 3) Panther Fusion SARS-CoV-2 (Hologic, Marlborough, MA), a high-throughput RT-qPCR method targeting open reading frame 1ab (ORF1ab); 4) Aptima SARS-CoV2 (Panther System, Hologic), a transcription mediated amplification method targeting ORF1ab; 5) GeneXpert Xpress SARS-CoV-2 (Cepheid, Sunnyvale, CA), a rapid RT-qPCR method targeting both *E* and *N* genes; 6) cobas Liat SARS-CoV-2 & Influenza A/B (Roche, Indianapolis, IN), a point-of-care RT-PCR method targeting ORF1ab and *N* gene; 7) e-Plex SARS-CoV-2 (Genmark, Carlsbad, CA), a rapid RT-PCR method targeting the *N* gene. All specimens testing positive for SARS-CoV-2 by NAAT with RT-qPCR C_t_ ≤ 30 or transcription-mediated amplification relative light units (RLU) ≥ 1,100 during this period were subject to multiplex allele-specific genotyping RT-qPCR. The small subset (288/4,049, 7.1%) of included specimens tested by rapid NAATs (Cepheid GeneXpert, Roche Liat, Genmark Eplex) did not have C_t_ values available and were included irrespective of viral load. All assays were conducted according to manufacturer and emergency authorization instructions.^7,8^

*Nucleic Acid Extraction*

Because residual eluate is not available for specimens originally tested on the Hologic or rapid NAAT (Cepheid GeneXpert, Roche Liat, Genmark Eplex) platforms, we re-extracted these genotyped samples from the original respiratory swab specimens on the same platform, provided there was sufficient material. Total nucleic acids were extracted from 300 µL viral transport media, universal transport media, or phosphate-buffered saline and eluted into 60 µL elution buffer (PerkinElmer Janus G3 Reformatter, Chemagic 360 nucleic acid extractor, and Chemagic Viral DNA/RNA 300 Kit).

All other specimens were genotyped from the residual diagnostic eluate.

**Supplementary Table 1.** Primer and Probe Oligonucleotide Sequences and Characteristics

| **Oligonucleotide** | **Sequence (5’ 🡪 3’)** | **5’ Modification** | **3’ Modification** | Tm^a^ Match (°C) | Tm^a^ Mismatch (°C) | **Sequence Conservation** | | | |
| --- | --- | --- | --- | --- | --- | --- | --- | --- | --- |
|  |  |  |  |  |  | **NCBI Pre-12/2020**  **(n=31,027)** | **GISAID B.1.427/B.1.429**  **(n=622)** | **GISAID B.1.1.7**  **(n=7,864)** | **GISAID B.1.351**  **(n=341)** |
| L452R_FWD | CTCTCTCAAAAGGTTTGAGATTAGACT | - | - | 62.7 | - | 99.6% (n=30,903) | 100.0% (n=622) | 99.9% (n=7,856) | 99.7% (n=340) |
| L452R_REV | CTTGATTCTAAGGTTGGTGGTAA | - | - | 60.5 | - | 99.0% (n=30,695) | 99.5% (n=619) | 99.8% (n=7,850) | 98.8% (n=337) |
| L452R_MT_HEX | CCTAAACAATCTATACCGGTAATT | HEX | BHQ | 58.7 | 51.6 | <0.1% (n=19) | 100.0% (n=622) | 0.0% (n=0) | 0.0% (n=0) |
| E484K_FWD | CTGAAATCTATCAGGCCGGTA | - | - | 61.2 | - | 99.4% (n=30,823) | 99.7% (n=620) | 99.7% (n=7,843) | 99.7% (n=340) |
| E484K_REV | GAAAGTACTACTACTCTGTATGG | - | - | 57.4 | - | 98.6% (n=30,584) | 99.7% (n=620) | 99.8% (n=7,850) | 98.5% (n=336) |
| E484K_MT_CY5 | CTTGTAATGGTGTTAAAGGTTT | CY5 | BHQ | 57.6 | 53.7 | <0.1% (n=13) | 0.0% (n=0) | <0.1% (n=1) | 99.7% (n=340) |
| N501Y_MT_FAM^b^ | TTTCCAACCCACTTATGGT | FAM | BHQ | 59.0 | 54.8 | 0.1% (n=38) | 0.0% (n=0) | 100.0% (n=7,861) | 99.7% (n=340) |
| N501_WT_CY3.5^b^ | TTTCCAACCCACTAATGGT | CY3.5 | BHQ | 59.0 | 56.8 | 99.2% (n=30,773) | 100.0% (n=622) | 0.0% (n=0) | 0.0% (n=0) |
| FWD, forward; REV, reverse; WT, wild-type; MT, mutant.  ^a^ Calculated using IDT OligoAnalyzer (<https://www.idtdna.com/pages/tools/oligoanalyzer>) using qPCR conditions (DNA, 0.2µM [oligonucleotide], 50mM [Na^+^], 3mM [Mg^2+^],0.8mM [dNTPs]. Mismatch Tm is for wild-type 🡪 mutant or mutant 🡪 wild-type nucleotide annealing.  ^b^ Anneals to E484K_FWD/REV amplicon downstream of E484K_MT_CY5. | | | | | | | | | |

**Supplementary Table 2.** Mutant Amplicon Controls

| **Oligonucleotide^a^** | **NC_045512 Polymorphism** | **Amplicon Length** | **Sequence (5’ 🡪 3’)** |
| --- | --- | --- | --- |
| ssDNA_L452R_MT | 22917 T>G | 79 | CTCTCTCAAAAGGTTTGAGATTAGACTTCCTAAACAATCTATACCGGTAATTATAATTACCACCAACCTTAGAATCAAG |
| ssDNA_del69-70_MT | del21765-70 | 113 | ACATTCAACTCAGGACTTGTTCTTACCTTTCTTTTCCAATGTTACTTGGTTCCATGCTATCTCTGGGACCAATGGTACTAAGAGGTTTGATAACCCTGTCCTACCATTTAATG |
| ssDNA_K417N_MT | 22813G>T | 101 | CATTTGTAATTAGAGGTGATGAAGTCAGACAAATCGCTCCAGGGCAAACTGGAAATATTGCTGATTATAATTATAAATTACCAGATGATTTTACAGGCTGC |
| ssDNA_E484K_MT | 23012G>A | 133 | CTGAAATCTATCAGGCCGGTAGCACACCTTGTAATGGTGTTAAAGGTTTTAATTGTTACTTTCCTTTACAATCATATGGTTTCCAACCCACTTATGGTGTTGGTTACCAACCATACAGAGTAGTAGTACTTTC |
| ssDNA_N501Y_MT | 23063A>T | 134 | GTTTTAATTGTTACTTTCCTTTACAATCATATGGTTTCCAACCCACTTATGGTGTTGGTTACCAACCATACAGAGTAGTAGTACTTTCTTTTGAACTTCTACATGCACCAGCAACTGTTTGTGGACCTAAAAAG |
| ssDNA_P681H_MT | 23604C>A | 99 | CAGGTATATGCGCTAGTTATCAGACTCAGACTAATTCTCATCGGCGGGCACGTAGTGTAGCTAGTCAATCCATCATTGCCTACACTATGTCACTTGGTG |
| MT, mutant.  ^a^ Individual ssDNA amplicons were pooled in equimolar ratios diluted to 10^4^ copies/µL each in Tris-EDTA buffer to create a mutant control. | | | |

**Supplementary Table 3.** Multiplex RT-PCR Reagents and Concentrations

| **Reagent** | **Stock Concentration** | **Volume (µL)** | **PCR Reaction Concentration** |
| --- | --- | --- | --- |
| L452R_FWD | 9000 nM | 1.0^a^ | 360 nM |
| L452R_REV | 9000 nM | - | 360 nM |
| E484K_FWD | 9000 nM | - | 360 nM |
| E484K_REV | 9000 nM | - | 360 nM |
| L452R_MT_HEX | 2000 nM | - | 80 nM |
| E484K_MT_CY5 | 2000 nM | - | 80 nM |
| N501Y_MT_FAM | 2000 nM | - | 80 nM |
| N501_WT_CY3.5 | 2000 nM | - | 80 nM |
| SuperScript™ III Platinum™ One-Step qRT-PCR Kit 2X Mix^b^ | 2X | 12.5 | 1X |
| SuperScript™ III Platinum™ One-Step qRT-PCR Kit *Taq* Mix^b^ | - | 0.5 | - |
| Nuclease-free Water | - | 6.0 | - |
| Template^c^ | - | 5.0 | - |
| **Total** | **-** | **25.0** | **-** |
| MT, mutant; WT, wild-type; nM, nanomolar.  ^a^ 1.0µL primer/probe mix at stock concentrations listed above.  ^b^ Catalog Numbers 11732-020 and 11732-088  ^c^ Wild-type TWIST whole-genome synthetic RNA control, pooled mutant amplicon control (Supplementary Table 2), or extracted nucleic acids from clinical upper respiratory swabs. | | | |

**Supplementary Table 4.** RT-qPCR Assay Interpretation and Reporting

| **Template** | **Instrument Result^a^** | | | | **Triage** | **Final Report** | | | | **Notes** |
| --- | --- | --- | --- | --- | --- | --- | --- | --- | --- | --- |
|  | **N501 (Cy3.5)** | **N501Y (FAM)** | **E484K (Cy5)** | **L452R (HEX)** | **Action** | **L452R Mutation** | **E484K Mutation** | **N501Y Mutation** | **Interpretative Comment^b^** | **Additional Guidance for Interpretation (Not Reported to Medical Record)** |
| Wild-type TWIST Control | C_t_<38 | ndet | ndet | ndet | Negative QC Passed | - | - | - | - | QC passed, proceed to interpretation of clinical specimens. |
| Mutant ssDNA Control | ndet | C_t_<38 | C_t_<38 | C_t_<38 | Positive QC Passed | - | - | - | - |  |
| Upper Respiratory Swab | C_t_ ≤40 | ndet | ndet | ndet | Report | Not Detected | Not Detected | Not Detected | 1 (below) | Suggests absence of variants listed below. |
| Upper Respiratory Swab | C_t_ ≤40 | ndet | C_t_ ≤40 | ndet | Report | Not Detected | Detected | Not Detected | 2 (below) | Presumptive P.2 variant vs. sporadic E484K mutation. |
| Upper Respiratory Swab | C_t_ ≤40 | ndet | ndet | C_t_ ≤40 | Report | Detected | Not Detected | Not Detected | 3 (below) | Presumptive B.1.427/B.1.429 variant vs. sporadic L452R mutation. |
| Upper Respiratory Swab | ndet | C_t_ ≤40 | C_t_ ≤40 | ndet | Report | Not Detected | Detected | Detected | 4 (below) | Presumptive B.1.351 variant vs. P.1 variant. |
| Upper Respiratory Swab | ndet | C_t_ ≤40 | ndet | ndet | Report | Not Detected | Not Detected | Detected | 5 (below) | Presumptive B.1.1.7 variant vs. sporadic N501Y mutation. |
| Upper Respiratory Swab | ndet | ndet | ndet | ndet | Repeat PCR | - | - | - | - | Extraction failure, setup failure, or viral load below limit of detection. Repeat PCR with 11µL eluate in same reaction volume (no water). If same result after repeat, report as “unable to genotype”. |
| Upper Respiratory Swab | C_t_ ≤40 | C_t_ ≤40 | C_t_ ≤40 | C_t_ ≤40 | Director Review | - | - | - | - | N501 and N501Y detected. Possible mixed infection, contamination, and/or nonspecific amplification. Medical director review. |
| Upper Respiratory Swab | C_t_ ≤40 | C_t_ ≤40 | C_t_ ≤40 | ndet | Director Review | - | - | - | - |  |
| Upper Respiratory Swab | C_t_ ≤40 | C_t_ ≤40 | ndet | C_t_ ≤40 | Director Review | - | - | - | - |  |
| Upper Respiratory Swab | C_t_ ≤40 | C_t_ ≤40 | ndet | ndet | Director Review | - | - | - | - |  |
| Upper Respiratory Swab | C_t_ ≤40 | ndet | C_t_ ≤40 | C_t_ ≤40 | Director Review | - | - | - | - | Mutations uncommonly observed together. Possible new variant, mixed infection, contamination, or nonspecific amplification. Medical director review. |
| Upper Respiratory Swab | ndet | C_t_ ≤40 | C_t_ ≤40 | C_t_ ≤40 | Director Review | - | - | - | - |  |
| Upper Respiratory Swab | ndet | C_t_ ≤40 | ndet | C_t_ ≤40 | Director Review | - | - | - | - |  |
| Upper Respiratory Swab | ndet | ndet | C_t_ ≤40 | C_t_ ≤40 | Director Review | - | - | - | - | N501 locus amplification failure. Medical director review. |
| Upper Respiratory Swab | ndet | ndet | C_t_ ≤40 | ndet | Director Review | - | - | - | - |  |
| Upper Respiratory Swab | ndet | ndet | ndet | C_t_ ≤40 | Director Review | - | - | - | - |  |
| C_t_, cycle threshold; ndet, not detected; RFU, relative fluorescence units.  ^a^ Channel thresholds: 500 RFU (FAM), 1000 RFU (HEX), 50 RFU (Cy3.5), 200 RFU (Cy5).  ^b^ Interpretative comments reported to medical record:  1. “These results do not rule out the presence of mutations other than L452R, E484K, and N501Y, or the possibility that L452R, N501Y, and/or E484K mutations are present below the assay lower limit of detection. These results should not be used as the sole basis for patient management decisions.”  2. “The presence of the E484K mutation without the N501Y mutation has been reported most commonly in SARS-CoV-2 lineages P.2, B.1.525, and B.1.526. It can also be seen sporadically, and in other lineages, and therefore cannot be used to definitively assign a variant/lineage/clade identity. Detection of E484K does not preclude the possibility of a mixed infection containing both mutant and wildtype SARS-CoV-2. These results do not rule out the presence of mutations other than L452R, E484K, and N501Y, or the possibility that N501Y and/or L452R are present at levels below the assay limit of detection. These results should not be used as the sole basis for patient management decisions.”  3. “The presence of the L452R mutation has been reported most commonly in SARS-CoV-2 lineages B.1.429 and B.1.427. It may be seen sporadically, and in other lineages, and therefore cannot be used to definitively assign a variant/lineage/clade identity. Detection of L452R does not preclude the possibility of a mixed infection containing both mutant and wildtype SARS-CoV-2. These results do not rule out the presence of mutations other than L452R, E484K, and N501Y, or the possibility that N501Y and/or E484K are present at levels below the assay limit of detection. These results should not be used as the sole basis for patient management decisions.”  4. “The presence of the E484K mutation and the N501Y mutation has been reported most commonly in SARS-CoV-2 lineages B.1.351 and P.1. They might also be seen sporadically and therefore cannot be used to definitively assign a variant/lineage/clade identity. Detection of these mutations does not preclude the possibility of a mixed infection containing both mutant and wildtype SARS-CoV-2. These results do not rule out the presence of mutations other than L452R, E484K, and N501Y, or the possibility that L452R is present at levels below the assay limit of detection. These results should not be used as the sole basis for patient management decisions.”  5. “The presence of N501Y mutation without the E484K mutation has been reported in a variety of SARS-CoV-2 lineages, most notably B.1.1.7. Detection of these mutations does not preclude the possibility of a mixed infection containing both mutant and wildtype SARS-CoV-2. These results do not rule out the presence of mutations other than L452R, E484K, and N501Y, or the possibility that E484K and/or L452 are present at levels below the assay limit of detection. These results should not be used as the sole basis for patient management decisions.” | | | | | | | | | | |

**Supplementary Table 5.** Analytical Performance: Lower Limit of Detection

| **Template and Concentration** | **Number of Detected Replicates** | | | |
| --- | --- | --- | --- | --- |
| **Mutant ssDNA Control** | **N501 Wild-Type**  **(Cy3.5)** | **N501Y Mutant**  **(FAM)** | **E484K Mutant**  **(Cy5)** | **L452R Mutant**  **(HEX)** |
| 1.0 copies / µL template | 0/20 | 8/20 | 7/20 | 17/20 |
| 5.0 copies / µL template | 0/20 | 9/20 | 11/20 | 20/20 |
| 10.0 copies / µL template | 0/20 | 14/20 | 16/20 | 20/20 |
| 100.0 copies / µL template | 0/20 | 20/20 | 20/20 | 20/20 |
| 95% LLOD (95% CI) - copies / µL template | (appropriate ndet^a^) | 23.6 (14.0-29.4) | 16.0 (11.1-40.3) | 1.5 (1.0-3.1) |
| 95% LLOD (95% CI) - copies / µL specimen^b^ | (appropriate ndet^a^) | 117.8 (70.0-147.2) | 79.9 (55.7-201.4) | 7.5 (5.0-15.6) |
| LLOD, lower limit of detection; CI, confidence interval; ndet, not detected.  ^a^ Appropriate absence of Cy3.5 amplification (N501 wild-type) in all replicates.  ^b^ Extrapolated from nucleic acid extraction protocol: 300 µL respiratory swab specimen extracted into 60 µL elution buffer. | | | | |

**Supplementary Table 6.** Analytical Performance: Linearity

| **Control Template** | **Concentration** | **Replicate C_t_ Values** | | | | **Coefficient of Variation (%)** | | | |
| --- | --- | --- | --- | --- | --- | --- | --- | --- | --- |
|  |  | **N501 Wild-Type**  **(Cy3.5)^a^** | **N501Y Mutant**  **(FAM)** | **E484K Mutant**  **(Cy5)** | **L452R Mutant**  **(HEX)** | **N501 Wild-Type**  **(Cy3.5)^a^** | **N501Y Mutant**  **(FAM)**^b^ | **E484K Mutant**  **(Cy5)**^b^ | **L452R Mutant**  **(HEX)**^b^ |
| **Mutant ssDNA** | 10^0^ copies / µL template | ndet x 3 | 42.61, 43.10, 43.70 | 42.95, 42.62, 42.56 | 43.35, 43.34, 43.34 | - | 0.40% | 0.20% | 0.10% |
|  | 10^1^ copies / µL template | ndet x 3 | 39.43, 40.40, 41.50 | 39.62, 40.24, 41.03 | 39.67, 40.02, 40.32 | - | 0.87% | 0.26% | 0.09% |
|  | 10^2^ copies / µL template | ndet x 3 | 37.31, 36.48, 36.06 | 37.18, 36.83, 36.50 | 36.25, 36.43, 36.25 | - | 0.38% | 1.14% | 0.15% |
|  | 10^3^ copies / µL template | ndet x 3 | 34.29, 34.16, 34.42 | 34.01, 33.88, 33.93 | 33.33, 33.20, 33.16 | - | 1.38% | 0.70% | 0.87% |
|  | 10^4^ copies / µL template | ndet x 3 | 30.09, 30.02, 30.12 | 30.25, 30.00, 30.27 | 29.54, 29.53, 29.57 | - | 9.00% | 5.21% | 1.47% |
|  | 10^5^ copies / µL template | ndet x 3 | 26.94, 26.72, 26.66 | 26.78, 26.70, 26.71 | 26.04, 26.05, 26.02 | - | 31.92% | 23.06% | 9.58% |
|  | 10^6^ copies / µL template | ndet x 3 | 23.14, 23.03, 22.98 | 23.25, 23.22, 23.17 | 22.65, 22.61, 22.64 | - | 98.99% | 32.26% | 10.58% |
| **Wild-type TWIST** | 10^4^ copies / µL template | 22.64 | ndet | ndet | ndet | - | - | - | - |
|  | 10^6^ copies / µL template | 27.33 | ndet | ndet | ndet | - | - | - | - |
| C_t_, cycle threshold.  ^a^ Appropriate absence of Cy3.5 amplification (N501 wild-type) in all replicates.  ^b^ Based on standard curves: N501Y-FAM: copies/µL template = 10^(12.888-0.295*C_t_); E484K-Cy5: copies/µL template = 10^(13.095-0.302*C_t_); L452R-HEX: copies/µL template = 10^(12.542-0.289*C_t_) | | | | | | | | | |

**Supplementary Table 7.** Whole Genome Next-Generation Sequencing Quality Measures for Subset of 229 Sequenced Specimens Genotyped by RT-qPCR

| **Sequencing Quality Measure** | **Median (Interquartile Range)** |
| --- | --- |
| Number of specimens | 229 |
| Diagnostic specimen NAAT C_t_ value | 17.9 (15.6 - 20.9) |
| Number of aligned reads | 664,369 (512,314 - 816,797) |
| Whole-genome coverage at 10X depth | 99.3% (99.0 - 99.4%) |
| Whole-genome mean coverage | 834.7 (689.5 - 911.4) |
| Spike protein coverage at 10X depth | >99.9% (99.9 - >99.9%) |
| Spike protein mean coverage | 829.2 (615.1 - 954.5) |
| NAAT, nucleic acid amplification test. | |

| **RT-qPCR Genotype Group** | **GISAID Accession Status** | **GISAID Accessions** |
| --- | --- | --- |
| L452, N501, E484 | Approved | >hCoV-19/USA/CA-Stanford-01_S03/2020\|EPI_ISL_1379719\|2020-12-10 |
|  |  | >hCoV-19/USA/CA-Stanford-01_S04/2020\|EPI_ISL_1424067\|2020-12-10 |
|  |  | >hCoV-19/USA/CA-Stanford-01_S05/2020\|EPI_ISL_1379720\|2020-12-10 |
|  |  | >hCoV-19/USA/CA-Stanford-01_S06/2020\|EPI_ISL_1379721\|2020-12-10 |
|  |  | >hCoV-19/USA/CA-Stanford-01_S16/2020\|EPI_ISL_1379722\|2020-12-15 |
|  |  | >hCoV-19/USA/CA-Stanford-01_S20/2020\|EPI_ISL_1379723\|2020-12-17 |
|  |  | >hCoV-19/USA/CA-Stanford-01_S21/2020\|EPI_ISL_1379724\|2020-12-19 |
|  |  | >hCoV-19/USA/CA-Stanford-01_S25/2020\|EPI_ISL_1424068\|2020-12-06 |
|  |  | >hCoV-19/USA/CA-Stanford-01_S26/2020\|EPI_ISL_1424069\|2020-12-06 |
|  |  | >hCoV-19/USA/CA-Stanford-01_S38/2020\|EPI_ISL_1424070\|2020-12-07 |
|  |  | >hCoV-19/USA/CA-Stanford-01_S39/2020\|EPI_ISL_1424071\|2020-12-07 |
|  |  | >hCoV-19/USA/CA-Stanford-02_S01/2021\|EPI_ISL_1379727\|2021-01-12 |
|  |  | >hCoV-19/USA/CA-Stanford-02_S02/2021\|EPI_ISL_1379728\|2021-01-12 |
|  |  | >hCoV-19/USA/CA-Stanford-02_S03/2021\|EPI_ISL_1379729\|2021-01-12 |
|  |  | >hCoV-19/USA/CA-Stanford-02_S04/2021\|EPI_ISL_1424072\|2021-01-12 |
|  |  | >hCoV-19/USA/CA-Stanford-02_S06/2020\|EPI_ISL_1379731\|2020-12-06 |
|  |  | >hCoV-19/USA/CA-Stanford-02_S07/2021\|EPI_ISL_1379732\|2021-01-12 |
|  |  | >hCoV-19/USA/CA-Stanford-02_S08/2020\|EPI_ISL_1379733\|2020-12-08 |
|  |  | >hCoV-19/USA/CA-Stanford-02_S10/2021\|EPI_ISL_1379734\|2021-01-12 |
|  |  | >hCoV-19/USA/CA-Stanford-02_S11/2021\|EPI_ISL_1379735\|2021-01-12 |
|  |  | >hCoV-19/USA/CA-Stanford-02_S15/2021\|EPI_ISL_1379737\|2021-01-12 |
|  |  | >hCoV-19/USA/CA-Stanford-02_S17/2021\|EPI_ISL_1379738\|2021-01-12 |
|  |  | >hCoV-19/USA/CA-Stanford-02_S21/2021\|EPI_ISL_1379739\|2021-01-12 |
|  |  | >hCoV-19/USA/CA-Stanford-02_S23/2021\|EPI_ISL_1379740\|2021-01-12 |
|  |  | >hCoV-19/USA/CA-Stanford-02_S24/2021\|EPI_ISL_1379741\|2021-01-12 |
|  |  | >hCoV-19/USA/CA-Stanford-02_S25/2021\|EPI_ISL_1379742\|2021-01-12 |
|  |  | >hCoV-19/USA/CA-Stanford-02_S27/2021\|EPI_ISL_1379743\|2021-01-12 |
|  |  | >hCoV-19/USA/CA-Stanford-02_S28/2021\|EPI_ISL_1379744\|2021-01-12 |
|  |  | >hCoV-19/USA/CA-Stanford-02_S29/2021\|EPI_ISL_1379745\|2021-01-12 |
|  |  | >hCoV-19/USA/CA-Stanford-02_S30/2021\|EPI_ISL_1379746\|2021-01-12 |
|  |  | >hCoV-19/USA/CA-Stanford-02_S32/2021\|EPI_ISL_1379747\|2021-01-12 |
|  |  | >hCoV-19/USA/CA-Stanford-02_S33/2021\|EPI_ISL_1379748\|2021-01-12 |
|  |  | >hCoV-19/USA/CA-Stanford-02_S37/2021\|EPI_ISL_1379749\|2021-01-12 |
|  |  | >hCoV-19/USA/CA-Stanford-02_S42/2021\|EPI_ISL_1379750\|2021-01-13 |
|  |  | >hCoV-19/USA/CA-Stanford-02_S45/2020\|EPI_ISL_1379751\|2020-12-13 |
|  |  | >hCoV-19/USA/CA-Stanford-02_S46/2021\|EPI_ISL_1424073\|2021-01-13 |
|  |  | >hCoV-19/USA/CA-Stanford-03_S08/2021\|EPI_ISL_1379756\|2021-01-05 |
|  |  | >hCoV-19/USA/CA-Stanford-03_S10/2021\|EPI_ISL_1379757\|2021-01-05 |
|  |  | >hCoV-19/USA/CA-Stanford-03_S11/2021\|EPI_ISL_1424076\|2021-01-05 |
|  |  | >hCoV-19/USA/CA-Stanford-03_S18/2021\|EPI_ISL_1424078\|2021-01-05 |
|  |  | >hCoV-19/USA/CA-Stanford-03_S21/2021\|EPI_ISL_1379763\|2021-01-05 |
|  |  | >hCoV-19/USA/CA-Stanford-03_S23/2021\|EPI_ISL_1379764\|2021-01-05 |
|  |  | >hCoV-19/USA/CA-Stanford-03_S24/2021\|EPI_ISL_1379765\|2021-01-05 |
|  |  | >hCoV-19/USA/CA-Stanford-03_S25/2021\|EPI_ISL_1379766\|2021-01-05 |
|  |  | >hCoV-19/USA/CA-Stanford-03_S27/2021\|EPI_ISL_1379768\|2021-01-05 |
|  |  | >hCoV-19/USA/CA-Stanford-03_S29/2021\|EPI_ISL_1379770\|2021-01-04 |
|  |  | >hCoV-19/USA/CA-Stanford-03_S31/2021\|EPI_ISL_1379771\|2021-01-02 |
|  |  | >hCoV-19/USA/CA-Stanford-03_S37/2021\|EPI_ISL_1424079\|2021-01-02 |
|  |  | >hCoV-19/USA/CA-Stanford-03_S38/2021\|EPI_ISL_1379774\|2021-01-02 |
|  |  | >hCoV-19/USA/CA-Stanford-03_S40/2021\|EPI_ISL_1424080\|2021-01-02 |
|  |  | >hCoV-19/USA/CA-Stanford-03_S42/2021\|EPI_ISL_1379776\|2021-01-02 |
|  |  | >hCoV-19/USA/CA-Stanford-03_S45/2021\|EPI_ISL_1364503\|2021-01-18 |
|  |  | >hCoV-19/USA/CA-Stanford-04_S05/2021\|EPI_ISL_1424081\|2021-01-23 |
|  |  | >hCoV-19/USA/CA-Stanford-04_S13/2021\|EPI_ISL_1379783\|2021-01-18 |
|  |  | >hCoV-19/USA/CA-Stanford-04_S14/2021\|EPI_ISL_1424083\|2021-01-24 |
|  |  | >hCoV-19/USA/CA-Stanford-04_S21/2021\|EPI_ISL_1379788\|2021-01-18 |
|  |  | >hCoV-19/USA/CA-Stanford-04_S23/2021\|EPI_ISL_1379789\|2021-01-19 |
|  |  | >hCoV-19/USA/CA-Stanford-04_S25/2021\|EPI_ISL_1379790\|2021-01-21 |
|  |  | >hCoV-19/USA/CA-Stanford-04_S35/2021\|EPI_ISL_1379796\|2021-01-20 |
|  |  | >hCoV-19/USA/CA-Stanford-04_S36/2021\|EPI_ISL_1424086\|2021-01-20 |
|  |  | >hCoV-19/USA/CA-Stanford-04_S38/2021\|EPI_ISL_1379798\|2021-01-20 |
|  |  | >hCoV-19/USA/CA-Stanford-04_S39/2021\|EPI_ISL_1379799\|2021-01-21 |
|  |  | >hCoV-19/USA/CA-Stanford-04_S40/2021\|EPI_ISL_1379800\|2021-01-21 |
|  |  | >hCoV-19/USA/CA-Stanford-04_S41/2021\|EPI_ISL_1379801\|2021-01-21 |
|  |  | >hCoV-19/USA/CA-Stanford-04_S43/2021\|EPI_ISL_1379803\|2021-01-22 |
|  |  | >hCoV-19/USA/CA-Stanford-04_S44/2021\|EPI_ISL_1379804\|2021-01-21 |
|  |  | >hCoV-19/USA/CA-Stanford-04_S46/2021\|EPI_ISL_1424087\|2021-01-21 |
|  |  | >hCoV-19/USA/CA-Stanford-05_S35/2021\|EPI_ISL_1379820\|2021-01-24 |
|  |  | >hCoV-19/USA/CA-Stanford-05_S36/2021\|EPI_ISL_1379821\|2021-01-23 |
|  |  | >hCoV-19/USA/CA-Stanford-05_S44/2021\|EPI_ISL_1379826\|2021-01-23 |
|  |  | >hCoV-19/USA/CA-Stanford-05_S47/2021\|EPI_ISL_1379827\|2021-01-20 |
|  |  | >hCoV-19/USA/CA-Stanford-06_S20/2021\|EPI_ISL_1424093\|2021-01-12 |
|  |  | >hCoV-19/USA/CA-Stanford-06_S39/2021\|EPI_ISL_1379848\|2021-01-29 |
|  |  | >hCoV-19/USA/CA-Stanford-06_S40/2021\|EPI_ISL_1379849\|2021-01-01 |
|  |  | >hCoV-19/USA/CA-Stanford-06_S45/2021\|EPI_ISL_1379853\|2021-01-31 |
|  |  | >hCoV-19/USA/CA-Stanford-07_S14/2021\|EPI_ISL_1379855\|2021-01-11 |
|  |  | >hCoV-19/USA/CA-Stanford-07_S27/2021\|EPI_ISL_1379856\|2021-01-07 |
|  |  | >hCoV-19/USA/CA-Stanford-07_S30/2021\|EPI_ISL_1379857\|2021-01-31 |
|  |  | >hCoV-19/USA/CA-Stanford-07_S42/2021\|EPI_ISL_1379866\|2021-01-29 |
|  |  | >hCoV-19/USA/CA-Stanford-07_S45/2021\|EPI_ISL_1379869\|2021-01-30 |
|  |  | >hCoV-19/USA/CA-Stanford-07_S46/2021\|EPI_ISL_1379870\|2021-01-29 |
|  |  | >hCoV-19/USA/CA-Stanford-08_S02/2021\|EPI_ISL_1379872\|2021-01-07 |
|  |  | >hCoV-19/USA/CA-Stanford-08_S04/2021\|EPI_ISL_1379874\|2021-01-23 |
|  |  | >hCoV-19/USA/CA-Stanford-08_S06/2021\|EPI_ISL_1379876\|2021-01-17 |
|  |  | >hCoV-19/USA/CA-Stanford-08_S07/2021\|EPI_ISL_1424103\|2021-01-23 |
|  |  | >hCoV-19/USA/CA-Stanford-08_S12/2021\|EPI_ISL_1379877\|2021-01-23 |
|  |  | >hCoV-19/USA/CA-Stanford-09_S13/2021\|EPI_ISL_1379884\|2021-01-22 |
|  |  | >hCoV-19/USA/CA-Stanford-09_S15/2021\|EPI_ISL_1424107\|2021-01-23 |
|  |  | >hCoV-19/USA/CA-Stanford-09_S16/2021\|EPI_ISL_1424108\|2021-01-23 |
|  |  | >hCoV-19/USA/CA-Stanford-09_S17/2021\|EPI_ISL_1424109\|2021-01-23 |
|  |  | >hCoV-19/USA/CA-Stanford-09_S18/2021\|EPI_ISL_1424110\|2021-01-23 |
|  |  | >hCoV-19/USA/CA-Stanford-09_S19/2021\|EPI_ISL_1424111\|2021-01-23 |
|  |  | >hCoV-19/USA/CA-Stanford-09_S23/2021\|EPI_ISL_1424112\|2021-01-24 |
|  |  | >hCoV-19/USA/CA-Stanford-09_S24/2021\|EPI_ISL_1379885\|2021-01-24 |
|  |  | >hCoV-19/USA/CA-Stanford-09_S25/2021\|EPI_ISL_1424113\|2021-01-24 |
|  |  | >hCoV-19/USA/CA-Stanford-09_S26/2021\|EPI_ISL_1379886\|2021-01-25 |
|  |  | >hCoV-19/USA/CA-Stanford-09_S30/2021\|EPI_ISL_1424114\|2021-01-25 |
|  |  | >hCoV-19/USA/CA-Stanford-09_S39/2021\|EPI_ISL_1424115\|2021-01-26 |
|  |  | >hCoV-19/USA/CA-Stanford-10_S23/2021\|EPI_ISL_1424126\|2021-01-27 |
|  |  | >hCoV-19/USA/CA-Stanford-10_S25/2021\|EPI_ISL_1424127\|2021-02-02 |
|  |  | >hCoV-19/USA/CA-Stanford-10_S38/2021\|EPI_ISL_1379887\|2021-02-03 |
|  |  | >hCoV-19/USA/CA-Stanford-10_S42/2021\|EPI_ISL_1379888\|2021-02-04 |
|  |  | >hCoV-19/USA/CA-Stanford-10_S43/2021\|EPI_ISL_1424129\|2021-02-05 |
| L452, N501, E484 | Pending Approval | >hCOV-19/USA/CA-Stanford-01_S27/2020\|pending\|2020-12-09 |
|  |  | >hCOV-19/USA/CA-Stanford-11_S08/2021\|pending\|2021-02-05 |
|  |  | >hCOV-19/USA/CA-Stanford-11_S10/2021\|pending\|2021-02-05 |
|  |  | >hCOV-19/USA/CA-Stanford-11_S12/2021\|pending\|2021-02-05 |
|  |  | >hCOV-19/USA/CA-Stanford-11_S13/2021\|pending\|2021-02-05 |
|  |  | >hCOV-19/USA/CA-Stanford-11_S15/2021\|pending\|2021-02-08 |
|  |  | >hCOV-19/USA/CA-Stanford-11_S16/2021\|pending\|2021-02-08 |
|  |  | >hCOV-19/USA/CA-Stanford-11_S17/2021\|pending\|2021-02-08 |
|  |  | >hCOV-19/USA/CA-Stanford-11_S18/2021\|pending\|2021-02-08 |
|  |  | >hCOV-19/USA/CA-Stanford-11_S20/2021\|pending\|2021-02-08 |
|  |  | >hCOV-19/USA/CA-Stanford-11_S22/2021\|pending\|2021-01-26 |
|  |  | >hCOV-19/USA/CA-Stanford-11_S23/2021\|pending\|2021-01-26 |
|  |  | >hCOV-19/USA/CA-Stanford-11_S30/2021\|pending\|2021-01-27 |
|  |  | >hCOV-19/USA/CA-Stanford-11_S33/2021\|pending\|2021-01-27 |
|  |  | >hCOV-19/USA/CA-Stanford-11_S36/2021\|pending\|2021-01-20 |
|  |  | >hCOV-19/USA/CA-Stanford-11_S39/2021\|pending\|2021-01-21 |
|  |  | >hCOV-19/USA/CA-Stanford-11_S40/2021\|pending\|2021-01-21 |
|  |  | >hCOV-19/USA/CA-Stanford-11_S43/2021\|pending\|2021-01-21 |
|  |  | >hCOV-19/USA/CA-Stanford-11_S44/2021\|pending\|2021-01-21 |
|  |  | >hCOV-19/USA/CA-Stanford-11_S45/2021\|pending\|2021-01-21 |
| L452R, N501, E484 | Approved | >hCoV-19/USA/CA-Stanford-01_S31/2020\|EPI_ISL_1379726\|2020-12-12 |
|  |  | >hCoV-19/USA/CA-Stanford-02_S12/2021\|EPI_ISL_1364488\|2021-01-12 |
|  |  | >hCoV-19/USA/CA-Stanford-02_S13/2021\|EPI_ISL_1379736\|2021-01-12 |
|  |  | >hCoV-19/USA/CA-Stanford-02_S14/2021\|EPI_ISL_1364489\|2021-01-12 |
|  |  | >hCoV-19/USA/CA-Stanford-02_S16/2021\|EPI_ISL_1364490\|2021-01-12 |
|  |  | >hCoV-19/USA/CA-Stanford-02_S18/2021\|EPI_ISL_1364491\|2021-01-12 |
|  |  | >hCoV-19/USA/CA-Stanford-02_S19/2021\|EPI_ISL_1364492\|2021-01-12 |
|  |  | >hCoV-19/USA/CA-Stanford-02_S20/2021\|EPI_ISL_1364493\|2021-01-12 |
|  |  | >hCoV-19/USA/CA-Stanford-02_S22/2021\|EPI_ISL_1364494\|2021-01-12 |
|  |  | >hCoV-19/USA/CA-Stanford-02_S34/2021\|EPI_ISL_1364495\|2021-01-12 |
|  |  | >hCoV-19/USA/CA-Stanford-02_S36/2021\|EPI_ISL_1364496\|2021-01-12 |
|  |  | >hCoV-19/USA/CA-Stanford-02_S39/2021\|EPI_ISL_1364497\|2021-01-13 |
|  |  | >hCoV-19/USA/CA-Stanford-02_S43/2021\|EPI_ISL_1364499\|2021-01-13 |
|  |  | >hCoV-19/USA/CA-Stanford-02_S47/2021\|EPI_ISL_1364500\|2021-01-13 |
|  |  | >hCoV-19/USA/CA-Stanford-03_S04/2021\|EPI_ISL_1379753\|2021-01-04 |
|  |  | >hCoV-19/USA/CA-Stanford-03_S06/2021\|EPI_ISL_1379754\|2021-01-05 |
|  |  | >hCoV-19/USA/CA-Stanford-03_S07/2021\|EPI_ISL_1379755\|2021-01-05 |
|  |  | >hCoV-19/USA/CA-Stanford-03_S09/2021\|EPI_ISL_1424075\|2021-01-05 |
|  |  | >hCoV-19/USA/CA-Stanford-03_S12/2021\|EPI_ISL_1379758\|2021-01-05 |
|  |  | >hCoV-19/USA/CA-Stanford-03_S13/2021\|EPI_ISL_1379759\|2021-01-05 |
|  |  | >hCoV-19/USA/CA-Stanford-03_S15/2021\|EPI_ISL_1379760\|2021-01-05 |
|  |  | >hCoV-19/USA/CA-Stanford-03_S16/2021\|EPI_ISL_1379761\|2021-01-04 |
|  |  | >hCoV-19/USA/CA-Stanford-03_S19/2021\|EPI_ISL_1379762\|2021-01-05 |
|  |  | >hCoV-19/USA/CA-Stanford-03_S26/2021\|EPI_ISL_1379767\|2021-01-05 |
|  |  | >hCoV-19/USA/CA-Stanford-03_S28/2021\|EPI_ISL_1379769\|2021-01-05 |
|  |  | >hCoV-19/USA/CA-Stanford-03_S34/2021\|EPI_ISL_1379772\|2021-01-02 |
|  |  | >hCoV-19/USA/CA-Stanford-03_S35/2021\|EPI_ISL_1379773\|2021-01-02 |
|  |  | >hCoV-19/USA/CA-Stanford-03_S39/2021\|EPI_ISL_1379775\|2021-01-02 |
|  |  | >hCoV-19/USA/CA-Stanford-03_S43/2021\|EPI_ISL_1379777\|2021-01-02 |
|  |  | >hCoV-19/USA/CA-Stanford-04_S04/2021\|EPI_ISL_1379780\|2021-01-24 |
|  |  | >hCoV-19/USA/CA-Stanford-04_S06/2021\|EPI_ISL_1379781\|2021-01-22 |
|  |  | >hCoV-19/USA/CA-Stanford-04_S11/2021\|EPI_ISL_1424082\|2021-01-18 |
|  |  | >hCoV-19/USA/CA-Stanford-04_S15/2021\|EPI_ISL_1379784\|2021-01-23 |
|  |  | >hCoV-19/USA/CA-Stanford-04_S16/2021\|EPI_ISL_1424084\|2021-01-18 |
|  |  | >hCoV-19/USA/CA-Stanford-04_S18/2021\|EPI_ISL_1379785\|2021-01-18 |
|  |  | >hCoV-19/USA/CA-Stanford-04_S19/2021\|EPI_ISL_1379786\|2021-01-18 |
|  |  | >hCoV-19/USA/CA-Stanford-04_S20/2021\|EPI_ISL_1379787\|2021-01-18 |
|  |  | >hCoV-19/USA/CA-Stanford-04_S27/2021\|EPI_ISL_1379792\|2021-01-19 |
|  |  | >hCoV-19/USA/CA-Stanford-04_S28/2021\|EPI_ISL_1424085\|2021-01-19 |
|  |  | >hCoV-19/USA/CA-Stanford-04_S29/2021\|EPI_ISL_1379793\|2021-01-19 |
|  |  | >hCoV-19/USA/CA-Stanford-04_S37/2021\|EPI_ISL_1379797\|2021-01-20 |
|  |  | >hCoV-19/USA/CA-Stanford-04_S42/2021\|EPI_ISL_1379802\|2021-01-21 |
|  |  | >hCoV-19/USA/CA-Stanford-05_S04/2021\|EPI_ISL_1379807\|2021-01-23 |
|  |  | >hCoV-19/USA/CA-Stanford-05_S13/2021\|EPI_ISL_1379809\|2021-01-22 |
|  |  | >hCoV-19/USA/CA-Stanford-05_S14/2021\|EPI_ISL_1379810\|2021-01-20 |
|  |  | >hCoV-19/USA/CA-Stanford-05_S16/2021\|EPI_ISL_1379811\|2021-01-20 |
|  |  | >hCoV-19/USA/CA-Stanford-05_S20/2021\|EPI_ISL_1379812\|2021-01-23 |
|  |  | >hCoV-19/USA/CA-Stanford-05_S29/2021\|EPI_ISL_1379816\|2021-01-22 |
|  |  | >hCoV-19/USA/CA-Stanford-05_S32/2021\|EPI_ISL_1379817\|2021-01-20 |
|  |  | >hCoV-19/USA/CA-Stanford-05_S37/2021\|EPI_ISL_1379822\|2021-01-22 |
|  |  | >hCoV-19/USA/CA-Stanford-05_S38/2021\|EPI_ISL_1379823\|2021-01-21 |
|  |  | >hCoV-19/USA/CA-Stanford-06_S06/2021\|EPI_ISL_1424090\|2021-01-31 |
|  |  | >hCoV-19/USA/CA-Stanford-06_S08/2021\|EPI_ISL_1379831\|2021-01-28 |
|  |  | >hCoV-19/USA/CA-Stanford-06_S14/2021\|EPI_ISL_1424091\|2021-01-31 |
|  |  | >hCoV-19/USA/CA-Stanford-06_S15/2021\|EPI_ISL_1379835\|2021-01-29 |
|  |  | >hCoV-19/USA/CA-Stanford-06_S16/2021\|EPI_ISL_1379836\|2021-01-28 |
|  |  | >hCoV-19/USA/CA-Stanford-06_S32/2021\|EPI_ISL_1379843\|2021-01-01 |
|  |  | >hCoV-19/USA/CA-Stanford-06_S38/2021\|EPI_ISL_1379847\|2021-01-31 |
|  |  | >hCoV-19/USA/CA-Stanford-06_S46/2021\|EPI_ISL_1424098\|2021-01-30 |
|  |  | >hCoV-19/USA/CA-Stanford-07_S31/2021\|EPI_ISL_1379858\|2021-01-30 |
|  |  | >hCoV-19/USA/CA-Stanford-07_S36/2021\|EPI_ISL_1379862\|2021-01-31 |
|  |  | >hCoV-19/USA/CA-Stanford-07_S37/2021\|EPI_ISL_1379863\|2021-01-30 |
|  |  | >hCoV-19/USA/CA-Stanford-07_S41/2021\|EPI_ISL_1379865\|2021-01-30 |
|  |  | >hCoV-19/USA/CA-Stanford-08_S35/2021\|EPI_ISL_1424105\|2021-01-31 |
|  |  | >hCoV-19/USA/CA-Stanford-08_S44/2021\|EPI_ISL_1379882\|2021-01-28 |
| L452R, N501, E484 | Pending Approval | >hCOV-19/USA/CA-Stanford-01_S19/2020\|pending\|2020-12-05 |
|  |  | >hCOV-19/USA/CA-Stanford-13_S41/2021\|pending\|2021-02-05 |
| L452, N501, E484K | Approved | >hCoV-19/USA/CA-Stanford-02_S40/2021\|EPI_ISL_1364498\|2021-01-13 |
|  |  | >hCoV-19/USA/CA-Stanford-04_S01/2021\|EPI_ISL_1364506\|2021-01-23 |
|  |  | >hCoV-19/USA/CA-Stanford-06_S07/2021\|EPI_ISL_1364507\|2021-01-29 |
|  |  | >hCoV-19/USA/CA-Stanford-07_S10/2021\|EPI_ISL_1424066\|2021-01-27 |
|  |  | >hCoV-19/USA/CA-Stanford-07_S13/2020\|EPI_ISL_1364508\|2020-12-31 |
|  |  | >hCoV-19/USA/CA-Stanford-07_S17/2021\|EPI_ISL_1364509\|2021-01-19 |
|  |  | >hCoV-19/USA/CA-Stanford-07_S18/2021\|EPI_ISL_1364510\|2021-01-19 |
|  |  | >hCoV-19/USA/CA-Stanford-07_S21/2021\|EPI_ISL_1364513\|2021-02-01 |
|  |  | >hCoV-19/USA/CA-Stanford-07_S22/2021\|EPI_ISL_1364514\|2021-02-01 |
|  |  | >hCoV-19/USA/CA-Stanford-07_S23/2021\|EPI_ISL_1364515\|2021-01-20 |
|  |  | >hCoV-19/USA/CA-Stanford-09_S01/2021\|EPI_ISL_1364516\|2021-01-27 |
|  |  | >hCoV-19/USA/CA-Stanford-09_S07/2021\|EPI_ISL_1364520\|2021-01-22 |
|  |  | >hCoV-19/USA/CA-Stanford-09_S10/2021\|EPI_ISL_1364523\|2021-02-11 |
| L452, N501, E484K | Pending Approval | >hCOV-19/USA/CA-Stanford-11_S06/2021\|pending\|2021-02-09 |
|  |  | >hCOV-19/USA/CA-Stanford-12_S41/2021\|pending\|2021-02-28 |
| L452, N501Y, E484 | Approved | >hCoV-19/USA/CA-Stanford-03_S44/2021\|EPI_ISL_1364502\|2021-01-15 |
|  |  | >hCoV-19/USA/CA-Stanford-03_S46/2021\|EPI_ISL_1364504\|2021-01-15 |
|  |  | >hCoV-19/USA/CA-Stanford-03_S47/2021\|EPI_ISL_1364505\|2021-01-18 |
|  |  | >hCoV-19/USA/CA-Stanford-07_S10/2021\|EPI_ISL_1424066\|2021-01-27 |
|  |  | >hCoV-19/USA/CA-Stanford-07_S20/2021\|EPI_ISL_1364512\|2021-01-19 |
|  |  | >hCoV-19/USA/CA-Stanford-09_S02/2021\|EPI_ISL_1364517\|2021-02-05 |
|  |  | >hCoV-19/USA/CA-Stanford-09_S03/2021\|EPI_ISL_1364518\|2021-01-26 |
|  |  | >hCoV-19/USA/CA-Stanford-09_S08/2021\|EPI_ISL_1364521\|2021-02-09 |
|  |  | >hCoV-19/USA/CA-Stanford-09_S09/2021\|EPI_ISL_1364522\|2021-02-11 |
|  |  | >hCoV-19/USA/CA-Stanford-10_S09/2021\|EPI_ISL_1424116\|2021-02-24 |
|  |  | >hCoV-19/USA/CA-Stanford-10_S10/2021\|EPI_ISL_1424117\|2021-02-24 |
|  |  | >hCoV-19/USA/CA-Stanford-10_S11/2021\|EPI_ISL_1424118\|2021-02-19 |
|  |  | >hCoV-19/USA/CA-Stanford-10_S12/2021\|EPI_ISL_1424119\|2021-02-21 |
|  |  | >hCoV-19/USA/CA-Stanford-10_S13/2021\|EPI_ISL_1424120\|2021-02-22 |
|  |  | >hCoV-19/USA/CA-Stanford-10_S14/2021\|EPI_ISL_1424121\|2021-02-18 |
|  |  | >hCoV-19/USA/CA-Stanford-10_S15/2021\|EPI_ISL_1424122\|2021-02-19 |
|  |  | >hCoV-19/USA/CA-Stanford-10_S16/2021\|EPI_ISL_1424123\|2021-02-19 |
|  |  | >hCoV-19/USA/CA-Stanford-10_S19/2021\|EPI_ISL_1424124\|2021-02-12 |
|  |  | >hCoV-19/USA/CA-Stanford-10_S21/2021\|EPI_ISL_1424125\|2021-02-26 |
| L452, N501Y, E484 | Pending Approval | >hCOV-19/USA/CA-Stanford-04_S22/2021\|pending\|2021-01-19 |
|  |  | >hCOV-19/USA/CA-Stanford-12_S37/2021\|pending\|2021-02-28 |
|  |  | >hCOV-19/USA/CA-Stanford-12_S38/2021\|pending\|2021-03-03 |
|  |  | >hCOV-19/USA/CA-Stanford-12_S46/2021\|pending\|2021-03-07 |
| L452, N501Y, E484K | Approved | >hCoV-19/USA/CA-05_S19_JAN/2021\|EPI_ISL_1335872\|2021-01-06 |
|  |  | >hCoV-19/USA/CA-07_S15_JAN/2021\|EPI_ISL_1335871\|2021-01-19 |
|  |  | >hCoV-19/USA/CA-Stanford-07_S10/2021\|EPI_ISL_1424066\|2021-01-27 |

**Supplementary Table 8.** GISAID Accession Identification Numbers for 229 Sequenced Specimens Genotyped by RT-qPCR

**Supplementary Figure 1.** Multiplex RT-qPCR linearity in replicates of three across seven orders of magnitude for the L452R (HEX), E484K (Cy5), and N501Y (FAM) allele-specific hydrolysis probes. Cycle threshold (C_t_) value is plotted against log10(copies/μL template) pooled ssDNA mutant amplicons.

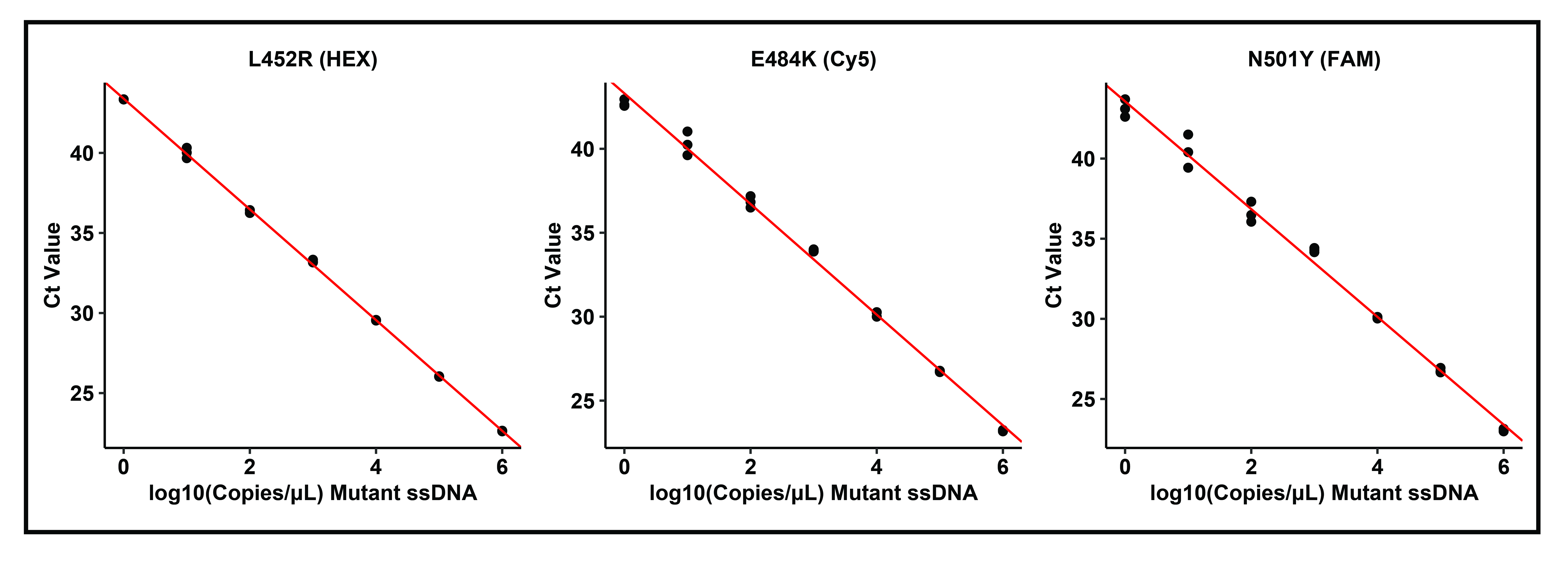
**Supplementary Figure 2.** Example multiplex genotyping RT-qPCR amplification curves for the four hydrolysis probes: L452R (HEX, yellow), E484K (Cy5, red), N501Y (FAM, green), and N501 (Cy3.5, orange). Horizontal lines represent each probe’s fluorescence threshold in the corresponding color. For specimens without N501Y mutations, N501 (Cy3.5) serves as an internal control in these known SARS-CoV-2 positive specimens. In addition to the ssDNA mutant control and TWIST wild-type whole-genome synthetic RNA control, the assay differentiates among at least five distinct genotypes depicted in this figure.

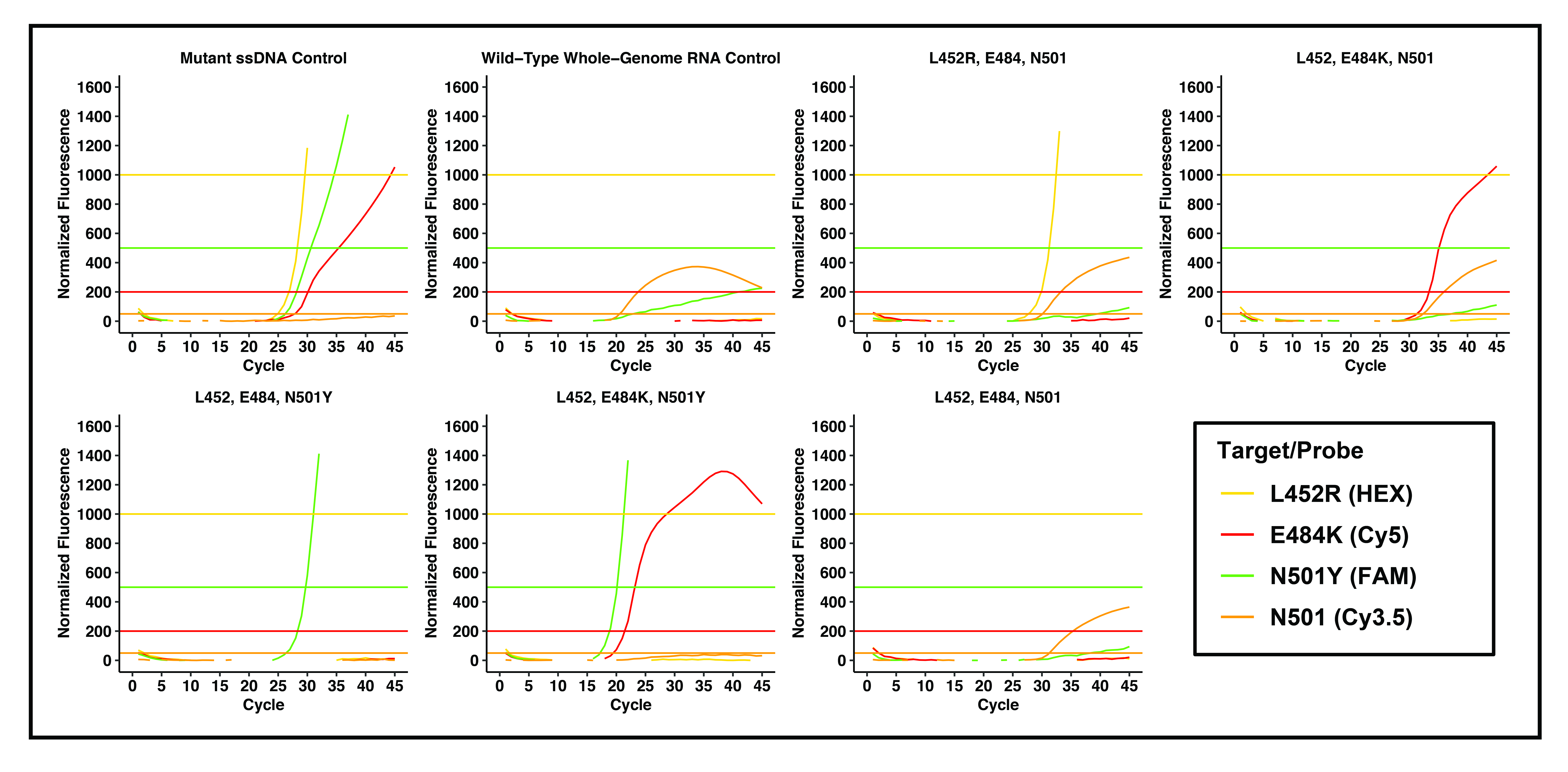
